# Cost-Utility Analysis of First-Line Olaparib plus Abiraterone for Metastatic Castration-Resistant Prostate Cancer in China after Volume-Based Procurement

**DOI:** 10.64898/2026.08.26.26361314

**Authors:** Hanqian Dai, Lijuan Jiang, Qiutao Gu, Chiyi Yao, Dandan Yu, Aidong Chao, Junwen Xie, Hui Xu, Ning Zhu, Bo Zhao, Liyuan Xu, Yingjie Du, Tianyi Wang, Shixuan Wang, Jiawei Pan, Anqi Yang, Jian Shi, Min Fan

## Abstract

**OBJECTIVE:** To evaluate the cost-utility and 5-year budget impact of first-line olaparib plus abiraterone versus abiraterone alone for metastatic castration-resistant prostate cancer (mCRPC) in China after the eleventh round of volume-based procurement (VBP). The intention-to-treat (ITT) population was assigned primary decision-analytic weight; the prespecified BRCA1/2-mutated (BRCAm) subgroup was a supporting analysis.

**METHODS:** A three-state partitioned survival model was built from the Chinese healthcare payer perspective over a 15-year horizon (5% discounting). ITT control-arm survival curves were reconstructed via the Guyot algorithm; the BRCAm control-arm survival curve was median-anchored to the published 23.0-month median. Utilities came from an mCRPC EQ-5D meta-analysis and a Chinese source, valued using the Chinese EQ-5D value set. Post-VBP prices were applied. A Markov model, probabilistic sensitivity analysis, and value-of-information analysis were performed.

**RESULTS:** The ITT incremental cost-effectiveness ratio (ICER) was CNY 40,198/ QALY (meta-analytic utilities) and CNY 43,721/QALY (Chinese utilities), below the CNY 287,247/QALY willingness-to-pay threshold (3× GDP). The BRCAm ICER was CNY 54,708/QALY and CNY 68,000/QALY, stable across the full OS hazard-ratio confidence interval and conservative survival-cap scenarios. All estimates remained cost-effective under a Markov structure (maximum CNY 120,558/QALY). Probabilistic cost-effectiveness exceeded 99.9%; population expected value of perfect information was zero. The 5-year budget impact was CNY 46.57 million.

**CONCLUSIONS:** At post-VBP prices, first-line olaparib plus abiraterone is cost-effective, robust across utility sources, survival distributions, and model structures. The BRCAm subgroup shows a hypothesis-consistent signal. These findings support National Reimbursement Drug List inclusion, particularly for the NMPA-approved BRCAm indication, contingent on adequate BRCA testing capacity.

**HIGHLIGHTS:**

- This is the first cost-utility analysis of first-line olaparib plus abiraterone for mCRPC in China at post-volume-based-procurement prices, addressing a gap left by prior pre-VBP Chinese evaluations.
- The combination is cost-effective across the intention-to-treat and BRCA1/2-mutated populations, robust to utility sources, survival distributions, model structures, and sensitivity analyses, with negligible decision uncertainty at the current willingness-to-pay threshold.
- Findings support National Reimbursement Drug List inclusion, particularly for the NMPA-approved BRCAm indication, contingent on strengthening BRCA testing capacity in tier-2 and tier-3 hospitals.

## 1. Introduction

Prostate cancer is the second most common cancer in men worldwide, with 1.47 million new diagnoses and 397,000 deaths in 2022.^1^ In China, the burden has risen rapidly as the population ages and screening expands. The national cancer registry recorded about 134,000 new cases and 48,000 deaths in 2022,^2–3^ and advanced-stage presentation is substantially more common than in Western populations.^3^ Metastatic castration-resistant prostate cancer (mCRPC) is the terminal stage of the disease. It remains the leading cause of prostate cancer mortality even after second-generation androgen-receptor pathway inhibitors were introduced.^4^ Around 10% of Western patients with mCRPC carry deleterious BRCA1 or BRCA2 mutations, and a comparable share of Chinese patients do (germline BRCA1/2 mutation prevalence 5.3-5.6% in unselected Chinese cohorts, rising with metastatic disease).^5–6^ This subgroup has a poor prognosis on androgen-receptor-directed monotherapy but responds strongly to poly(ADP-ribose) polymerase (PARP) inhibition.^7–8^

Three phase 3 mCRPC trials investigated first-line PARP inhibitor combinations. PROpel randomised 796 unselected patients to abiraterone plus olaparib versus abiraterone plus placebo (rPFS 24.8 vs 16.6 months, HR 0.66, 95% CI 0.54-0.81; final OS 42.1 vs 34.7 months, HR 0.81, 0.67-1.00).^9,7^ The effect was much larger in its prespecified BRCA1/2-mutated (BRCAm) subgroup (85 patients: 47 olaparib plus abiraterone, 38 placebo plus abiraterone; 10.9% of the trial, identified by aggregated tumour tissue and circulating tumour DNA testing). In this subgroup the rPFS HR was 0.23 (95% CI 0.12-0.43, investigator-assessed; blinded independent central-review results were similar). At the final descriptive OS analysis the OS HR was 0.29 (0.14-0.56), with control-arm median OS of 23.0 months and intervention-arm median OS not reached (13/47 vs 25/38 deaths).^9,7^ An earlier data cut-off had reported an immature interim OS estimate (HR 0.30), but the more mature final value superseded it; we use the final value throughout this analysis. This BRCAm subgroup is distinct from the broader homologous-recombination-repair-mutated (HRRm) subgroup (n=226), whose final OS HR was 0.66 (0.45-0.95)^7^; the present analysis models the BRCA1/2-confirmed population.

A subsequent FDA pooled analysis of four PARP-inhibitor trials confirmed the largest benefit in BRCA1/2-altered patients.^10,8^ MAGNITUDE^11–12^ and TALAPRO-2^13–15^ similarly established BRCAm-selected first-line indications for niraparib plus abiraterone and talazoparib plus enzalutamide.

China has changed the regulatory and pricing environment for this indication markedly. The US Food and Drug Administration approved olaparib plus abiraterone for the BRCAm subgroup in May 2023^8^; China’s National Medical Products Administration (NMPA) followed on 31 July 2025, based on the PROpel BRCAm subgroup and Chinese cohort trend-consistency data.^16^ In parallel, the eleventh round of national volume-based procurement (VBP) cut the olaparib price by more than 98%, from about CNY 12,240 to CNY 188.7 per 28-day cycle at 300 mg twice daily.^17^ Abiraterone had already fallen to about CNY 717.6 per cycle through earlier rounds.^18^ The reference willingness-to-pay (WTP) threshold has also been updated to CNY 287,247/QALY, three times the 2024 Chinese per-capita GDP of CNY 95,749.^19^ In the later-line landscape, the November 2025 NMPA approval of lutetium-177 vipivotide tetraxetan (Pluvicto) for PSMA-positive mCRPC after an androgen-receptor pathway inhibitor introduces a costly post-progression option.^20–21^ This is directly relevant to any first-line reimbursement decision. Despite these clinical and pricing changes, the only prior Chinese cost-utility analysis (CUA) of olaparib evaluated monotherapy in the post-progression PROfound population. That study used pre-VBP prices and pre-dated the current pharmacoeconomic evaluation guideline; it reported an ICER of CNY 392,728/QALY and concluded that olaparib was not cost-effective in China.^22–23^ No study has quantified the economic value of first-line olaparib plus abiraterone under the current pricing environment.

This study assessed the cost-utility of first-line olaparib plus abiraterone versus abiraterone alone for mCRPC in China. We assigned the primary decision-analytic weight to the intention-to-treat (ITT) population, which is anchored to the larger PROpel evidence base (N = 796) and is comparable with regulatory decisions in unselected first-line populations.^11–15^ We analysed the prespecified BRCA1/2-mutated (BRCAm) subgroup, which matches the current NMPA indication, as a supporting precision-medicine scenario. We built a three-state partitioned survival model (PSM) from the PROpel survival data. For the ITT population we used digitally reconstructed pseudo-individual patient data (Guyot algorithm),^24^ selecting lognormal distributions on information criteria and carrying generalised gamma (GG) and log-logistic distributions as parametric alternatives. For the BRCAm subgroup we used a median-anchored, maturity-constrained parametric control-arm OS curve, because its published Kaplan-Meier plot is too sparse for reliable digitisation. The design combined dual utility sourcing (a pooled EQ-5D meta-analysis in mCRPC^25^ and Chinese first-line source utilities^22^), a discrete-time Markov cohort model as a co-primary structural comparator,^26–28^ a 5-year budget impact analysis, and an accompanying value of information (VOI) analysis. Alongside this design, the analysis introduces four methodological refinements relative to prior Chinese work in the same disease area. First, ITT pseudo-IPD reconstruction from the PROpel Number-at-Risk table uses distribution selection guided by AIC and BIC, consistent with NICE Decision Support Unit Technical Support Document 14.^29–30^ Second, we quantify PSM PD-clipping frequency explicitly for each scenario (PD-clipping is the artefact by which the partitioned-survival structure lets modelled deaths occur only after progression, clipping post-progression occupancy when the PFS curve rises above the OS curve). Third, we apply a conservative HR_OS = HR_PFS bound as a sensitivity analysis on the ITT ICER. Fourth, we compute expected value of partial perfect information (EVPPI) under three independent metamodels (random-forest in-sample, random-forest out-of-bag, LinearGAM).^31–33^ Reporting followed the Consolidated Health Economics Evaluation Reporting Standards (CHEERS) 2022.^34^ Section 3 answers the headline question, Section 4 tests the robustness of the modelling assumptions, and Section 5 sets the base case in policy context; readers looking only for the headline may go directly to §3.1 and §3.4.

## 2. Methods

### 2.1 Study design, comparators and analytic overview

This was a model-based cost-utility analysis (CUA) conducted from the Chinese healthcare payer (National Reimbursement Drug List, NRDL) perspective and reported according to CHEERS 2022.^34^ We analysed two populations in parallel. We assigned the primary decision-analytic weight to the intention-to-treat (ITT) population of men with first-line mCRPC, because it rests on the larger PROpel evidence base (N=796) and mirrors regulatory decisions on niraparib plus abiraterone^11–12^ and talazoparib plus enzalutamide^13–15^ in unselected or broader homologous recombination repair (HRR)-mutated populations. We analysed the prespecified BRCAm subgroup — adult men with BRCA1/2-mutated mCRPC eligible for first-line combination therapy, the population matching the current NMPA indication^16^ — as a supporting precision-medicine scenario. The intervention was olaparib 300 mg twice daily plus abiraterone 1,000 mg once daily plus prednisone 5 mg twice daily; the comparator was placebo plus abiraterone 1,000 mg once daily plus prednisone 5 mg twice daily, matching the PROpel control arm^9^ and current Chinese practice in chemotherapy-ineligible mCRPC.^16^ The primary analytic engine was a three-state partitioned survival model (PSM), and we ran a discrete-time Markov cohort model in parallel as a structural comparator. We reported four base cases by crossing two populations (BRCAm subgroup, ITT) with two utility sources (Castro 2024 pooled EQ-5D meta-analysis; Xu 2022 Chinese utilities). Four additional methodological sensitivity analyses (pseudo-IPD reconstruction, per-scenario PD-clipping, HR bound, PD-cost multiplier) accompany the base analyses (§4.1-§4.4).

### 2.2 Health economic analysis plan

We did not conduct this analysis under a prospectively registered Health Economic Analysis Plan (HEAP), which we acknowledge as a limitation of a Chinese national reimbursement-relevant CUA.^35^ We finalised the scope, comparators, perspective, time horizon, discount rate, base cases and outcome measures before running the deterministic and probabilistic analyses. We document the analytic decisions taken after the initial run (pseudo-IPD reconstruction, distributional alternatives, EVPPI metamodel checks) with their rationales in §4.1-§4.4 and §5.4. Consistent with the pre-planned versus post-hoc distinction recommended for transparent reporting, we pre-planned the base cases, comparators, perspective, horizon, discount rate, primary utility source, WTP threshold and the PSM/Markov structural comparison before any analysis was run. The following were run post hoc and are labelled as such throughout: the Guyot pseudo-IPD reconstruction (§4.1), the generalised-gamma and log-logistic distributional alternatives (§3.2), the non-proportional-hazards AFT sensitivity path (§3.5), the two-transition Markov calibration (§3.2), the per-scenario PD-clipping quantification (§4.2), the multi-seed and probabilistic budget-impact analyses (§3.6), the MAGNITUDE external-HR scenario (§5.3) and the EVPPI metamodel robustness checks (§2.6). Ethics approval. This study used only previously published, aggregate, de-identified trial data and reconstructed pseudo-individual patient data derived from published Kaplan-Meier curves; we accessed no individual patient records and contacted no human participants. Institutional review board approval and informed consent were therefore not required, consistent with CHEERS 2022 Item 5 and standard practice for secondary analyses of published data.

### 2.3 Patient and public involvement

We engaged no formal patient and public involvement mechanism during the design or execution of this analysis.^34^ The closest proxy for Chinese-patient input is the Xu et al. 2022 EQ-5D-3L utility source,^22^ which sampled 30 Chinese mCRPC patients receiving PARP inhibitor therapy and mapped their responses to the Chinese EQ-5D-3L value set^36^; we retain these values as a Chinese-anchored scenario. The Castro et al. 2024 pooled EQ-5D meta-analysis utilities^25^ used in the primary base case were derived from mCRPC patients outside China and do not represent Chinese-patient preferences directly. The two utility sources produce BRCAm ICERs of CNY 54,708/QALY (Castro) and 68,000/QALY (Xu), a 24% swing that is material to the cost-effectiveness interpretation. We judge the Castro 2024 meta-analytic values the more appropriate primary source for three reasons. First, they pool 45 mCRPC studies (12 meta-analysable) and therefore carry far greater precision than the Xu 2022 sample of 30 Chinese patients. Second, they report a genuine first-line utility (0.79), whereas Xu’s values are second-line-derived and, when applied to a first-line population, bias the ICER upward. Third, both sources are mapped onto the same Chinese EQ-5D value set,^36^, so the comparison isolates the population and line-of-therapy difference rather than a valuation-tariff difference. We therefore interpret the Xu-based ICER as a conservative Chinese-anchored upper bound rather than as an equally weighted alternative, and the 24% swing as a one-directional conservatism rather than symmetric uncertainty. Chinese first-line disease-state EQ-5D data would resolve the residual gap, and we identify them as a research priority. Future Chinese CUAs of first-line PARP inhibitor combinations should elicit Chinese first-line disease-state utilities, especially because the VOI analysis (§3.4) points to PFS management resource use, not survival or utility parameters, as the principal contributor to residual decision uncertainty in the BRCAm setting.

### 2.4 Model structure

The three-state PSM comprised progression-free (PFS), progressed disease (PD) and death. State occupancy in each 28-day cycle was derived from the parametric survival functions using the standard PSM identities:

- P(PFS)(t) = S_PFS(t)
- P(Death)(t) = 1 − S_OS(t)
- P(PD)(t) = S_OS(t) − S_PFS(t)

This construction maps the two trial-observed survival endpoints (rPFS and OS) directly to state membership.^26,37^ The model horizon was 15 years, sufficient to capture more than 99% of the expected discounted survival benefit; the cycle length of 28 days matched PROpel treatment scheduling, and we applied a half-cycle correction.^28^ We constructed a parallel discrete-time Markov cohort model with the same three states and the same costs and utilities as a structural comparator.^27^ In the Markov model, we calibrated the PFS→PD and PD→Death transition probabilities to each arm’s median PFS and median OS-PFS gap respectively and held them constant across cycles.

### 2.5 Clinical data and survival curves

Individual-patient-level PROpel data are not publicly available, and the two model populations required different survival-curve strategies because their published Kaplan-Meier data differ greatly in maturity. For the intention-to-treat (ITT) population, we digitised the published control-arm rPFS and OS Kaplan-Meier curves^7^ (N=397 placebo; 205 OS events) in WebPlotDigitizer and regenerated pseudo-individual patient data (pseudo-IPD) with the Guyot algorithm,^24^ following the National Institute for Health and Care Excellence Decision Support Unit Technical Support Document 14 recommendation to use pseudo-IPD rather than assumed parametric shapes.^29–30^ We fitted six candidate parametric distributions (exponential, Weibull, lognormal, log-logistic, Gompertz, gamma) to each reconstructed ITT dataset by maximum likelihood, guiding selection by Akaike Information Criterion (AIC), Bayesian Information Criterion (BIC) and visual fit. We then re-fit three accelerated failure time (AFT) distributions (lognormal, generalised gamma (GG), log-logistic) to the same ITT pseudo-IPD in flexsurv v2.3. We report the ICER envelope across them in §3.5 and Figure 6.

For the BRCA1/2-mutated (BRCAm) subgroup we did not use this Guyot-plus-multi-distribution approach, because the published BRCAm control-arm survival data are too sparse for reliable digitisation or parametric-distribution selection. The BRCAm placebo arm comprises only 38 patients with 25 OS events, and the corresponding Kaplan-Meier curve is a coarse step function that cannot support Guyot reconstruction or discrimination among competing parametric tails without spurious precision (NICE DSU TSD 14).^29^ Instead, we constructed the BRCAm control-arm OS curve with a median-anchored, maturity-constrained parametric approach. We anchored a lognormal curve to the published BRCAm control-arm median OS of 23.0 months^9,7^ (log-scale location μ = ln 23.0 = 3.1355). We then solved the scale parameter σ (the log-scale standard deviation of the lognormal survivor function) so that the survivor function reproduced the observed BRCAm control-arm mortality at the PROpel data cut-off (S(36.6 months) = 1 − 25/38 = 0.342, giving σ = 1.1422). This single, published-anchor-consistent curve is the BRCAm base case. The three Guyot-fitted parametric alternatives (lognormal/GG/ log-logistic) are therefore not applicable to the BRCAm subgroup; we instead quantify parametric uncertainty for BRCAm probabilistically through the PSA (§3.4, §3.5) rather than through a distributional matrix. We chose the lognormal form for the BRCAm anchor for clinical-plausibility reasons that also motivate its retention for the ITT base case. The lognormal hazard is single-peaked and declines in the tail, qualitatively consistent with mCRPC OS follow-up from PROfound^38–39^ and TALAPRO-2^15^ and with the biological expectation that early post-progression mortality peaks and then falls as susceptible patients leave the risk set. For the ITT population we retained the lognormal as base case even though it is not the AIC-optimal fit on the ITT Guyot pseudo-IPD (ITT Δ AIC 17.0 vs the best-fitting distribution). Observed-range fit was similar across candidates, while their consequential differences arose primarily in the unobserved 5-15 year extrapolation tail, a region a data-driven AIC preference cannot arbitrate. We therefore disclose the full ITT distributional ICER envelope in Figure 6 and Supplementary Table S2 rather than reducing it to a single point estimate. §5.5(9) gives a fuller version of this defence. So that the reader is not dependent on the lognormal choice for ITT, we co-report the generalised gamma and log-logistic ITT ICERs as reference values (ITT × Castro CNY 40,198 lognormal, 46,955 generalised gamma, 36,765 log-logistic). Because the generalised gamma is the AIC-preferred ITT distribution, we highlight its ITT × Castro ICER of CNY 46,955/QALY (+16.8% vs lognormal) as the reference upper case; at 16.3% of the WTP threshold it does not alter any cost-effectiveness conclusion.

The base-case control-arm lognormal parameters were as follows. The ITT parameters were estimated by maximum likelihood on the Guyot pseudo-IPD: ITT rPFS μ=2.9865, σ=1.0453 (fitted median 19.82 months); ITT OS μ=3.5555, σ=1.1179 (fitted median 35.01 months). The BRCAm control-arm OS parameters were set by the median-anchored, maturity-constrained construction described above: μ=3.1355, σ=1.1422 (median 23.0 months, reproducing the published BRCAm control mortality of 25/38 = 65.8% at the 36.6-month cut-off), with the BRCAm control-arm rPFS anchored to its published median of 8.0 months (μ=2.0794). Because the model applies proportional-hazards (PH) scaling S_int(t) = S_ctrl(t)^HR at the model level, intervention-arm curves are derived from the control-arm parameters combined with the reported hazard ratios. For the BRCAm subgroup these are rPFS HR 0.23 (95% CI 0.12-0.43, investigator-assessed; blinded independent central-review results were similar) and final OS HR 0.29 (0.14-0.56) ^9,7^; for ITT, rPFS HR 0.66 (0.54-0.81) and OS HR 0.81 (0.67-1.00) ^9,7^. Under PH scaling, the median of the BRCAm intervention-arm OS curve is approximately 105.2 months; by contrast, 23.0/0.29 = 79.3 months is the naive median-ratio/AFT anchor used only in the supplementary AFT and Markov calibration scenarios. The PH-scaled curve is consistent with the published intervention-arm OS being not reached, and the modelled intervention-arm mortality at the 36.6-month cut-off (26.7%) closely matches the observed 13/47 = 27.7%. Log-cumulative-hazard plots for the reconstructed ITT rPFS and OS (Figure 1) were approximately parallel for rPFS (slope ratio 1.02) and mildly divergent for OS (slope ratio 1.19), consistent with the modest OS proportional-hazards deviation reported for maintenance PARP inhibitors ^9^; we therefore added a non-PH AFT sensitivity path (§3.5; post hoc, §2.2). Because the OS log-cumulative-hazard slope ratio (1.19) exceeded the pre-specified 1.15 threshold for departure from proportional hazards, the BRCAm base-case ICER is reported under BOTH structures. On the median-anchored BRCAm control curve with the true OS HR 0.29, the BRCAm × Castro base case has a mean PSA ICER of CNY 56,461/QALY under the proportional-hazards structure (Path A) and CNY 56,172/QALY under the accelerated-failure-time non-PH structure (Path B). This is a difference of only 0.5%. Although the two structures imply different intervention-arm OS medians (approximately 105.2 months under PH scaling and 79.3 months under the prespecified AFT shift), their mean ICERs differ by only 0.5%. The deterministic PH base of this analysis reproduces the primary BRCAm × Castro base-case ICER of CNY 54,708/QALY exactly. This confirms that the headline result does not depend on the proportional-hazards assumption (full paths in Supplementary Table S2 Panel A and §3.5).

**Figure 1.**
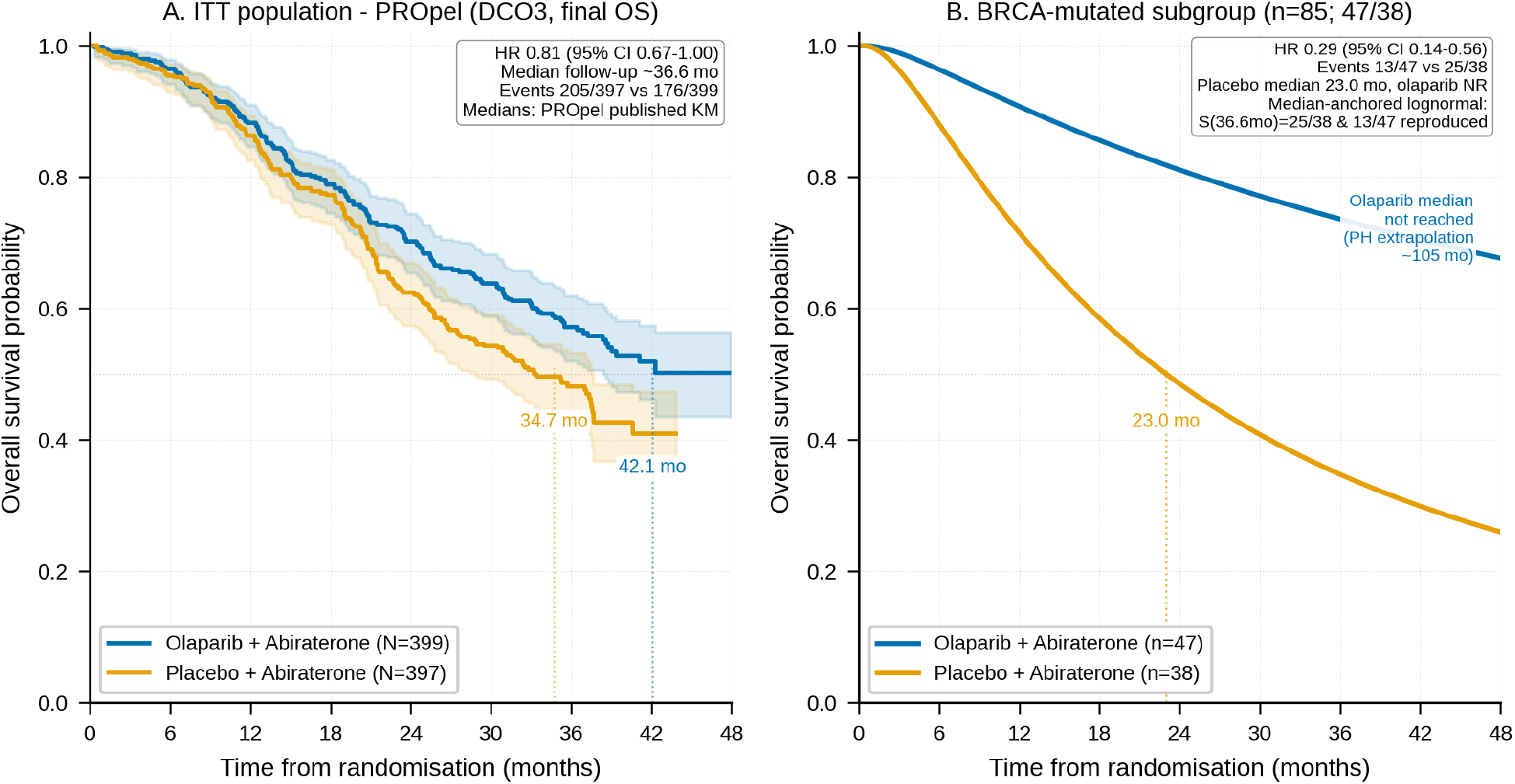
Overall survival curves for the two model populations, constructed by the two methods their differing data maturity requires (§2.5). Panel A: ITT population overall survival Kaplan-Meier curves reconstructed from the PROpel Number-at-Risk table (Saad 2023, Lancet Oncol) using the Guyot algorithm (N=796; N=397 placebo, N=399 olaparib+abiraterone; total 381 events, 205 placebo/176 intervention, within one event of the published 205/176 total); confidence bands are 95% pointwise Greenwood intervals. Panel B: BRCA1/2-confirmed BRCAm subgroup (n=85; 38 placebo, 47 olaparib+abiraterone) — because the BRCAm placebo arm (38 patients, 25 OS events) is too sparse for reliable Guyot digitisation, the control-arm OS curve is a median-anchored, maturity-constrained lognormal fixed to the published BRCAm control median OS of 23.0 months (meanlog μ = ln 23.0 = 3.1355 log-months, sdlog σ = 1.1422, reproducing the observed 25/38 = 65.8% control mortality at the 36.6-month data cut-off); the intervention-arm curve is derived by proportional-hazards scaling with the true BRCAm OS HR of 0.29, giving a modelled intervention OS median of ∼105 months (published intervention median not reached). The ITT Guyot-reconstructed pseudo-IPD are used as the primary input for ITT parametric survival fitting.

### 2.6 Utilities, costs and adverse events

We compared two utility sources. The base case used the pooled EQ-5D index values reported by Castro et al. (2024), a systematic literature review and meta-analysis of 45 mCRPC publications with 12 studies feasible for meta-analysis: 0.79 (95% CI 0.75-0.84) for first-line treatment and 0.69 (0.67-0.71) for second-line-and-later treatment.^25^ Following standard mCRPC modelling convention, we mapped the first-line pooled utility to the PFS state (dominated by patients on first-line treatment before radiographic progression) and the second-line-and-later utility to the PD state. A scenario applied the Chinese first-line utilities of Xu et al. (2022): PFS 0.617, PD 0.37,^22^ both derived from a PROfound-based Chinese olaparib monotherapy CUA in a second-line-and-later population and adopted in three subsequent Chinese prostate cancer CUAs^40–42^. We retain their use as first-line utilities in the present analysis as a conservative scenario. Applying second-line utilities (Xu 0.617/0.37) to a first-line population understates first-line health-related quality of life. It therefore reduces incremental QALYs on the same incremental-cost trajectory, biasing the ICER upward relative to a first-line-derived utility set. §5.4 also acknowledges this mismatch. We applied adverse-event disutilities as one-time first-cycle decrements (anaemia −0.119; nausea −0.21).^22^

We expressed costs in 2025 Chinese Yuan (CNY) from the healthcare payer perspective. For international readers and CHEERS 2022 Item 15, an approximate USD equivalent uses the People’s Bank of China central parity rate on 30 June 2025 (1 USD ≈ 7.16 CNY): the base-case BRCAm × Castro ICER of CNY 54,708/QALY corresponds to about USD 7,600/QALY, and the WTP threshold of CNY 287,247/ QALY to about USD 40,100/QALY. Drug acquisition costs used post-VBP prices. We priced olaparib (150 mg × 56 tablets) at CNY 188.7 per box from the eleventh round of national VBP,^17^ yielding CNY 188.7 per 28-day cycle at 300 mg twice daily. We priced abiraterone acetate (0.25 g × 120 tablets) at CNY 717.6 per box under the Shandong Province VBP continuation,^18^ yielding CNY 717.6 per 28-day cycle at 1,000 mg once daily. Prednisone was CNY 3.78 per cycle.^22^ Non-drug PFS management cost was CNY 2,595.75 per cycle (laboratory tests, computed tomography, prostate-specific antigen, bone imaging, nursing fees, routine follow-up); post-progression management cost was CNY 5,831.53 per cycle, dominated by docetaxel chemotherapy applied to 50% of patients as a conservative assumption plus continuing supportive care.^22^ We applied adverse-event costs one-time in the first cycle (anaemia CNY 3,893.05; nausea CNY 467.49).^22^

We took adverse-event rates directly from the PROpel safety population (n=398 intervention, n=396 control).^7^ Grade ≥3 anaemia rates were 64/398 (16.1%) in the intervention arm and 13/396 (3.3%) in the control arm; any-grade nausea rates were 122/398 (30.7%) versus 57/396 (14.4%).^7^ We selected any-grade nausea rates because the Xu 2022 nausea disutility of −0.21 is calibrated to symptomatic nausea rather than grade ≥3-only nausea.^22^ Using the grade ≥3-only rate (0.3% in the intervention arm) would markedly under-count the symptomatic burden the disutility represents. Sensitivity checks confirm insensitivity: the between-arm AE-related QALY differential is 0.001-0.003 QALYs, so replacing any-grade with grade ≥3 nausea changed base-case ICERs by less than 0.6%.

### 2.7 Sensitivity analyses framework

The primary outcome was the ICER expressed as CNY per QALY gained, compared with the WTP threshold of three times the 2024 Chinese per-capita GDP (CNY 95,749),^19^ yielding CNY 287,247/QALY, consistent with the Chinese Guidelines for Pharmacoeconomic Evaluations 2020.^23,43^ We discounted both costs and quality-adjusted life-years (QALYs)^44^ at 5% annually.^23^ The sensitivity architecture comprised five layers. Layer (i) was deterministic one-way sensitivity of the base-case ICER to key parameters (Figure 2 tornado). Layer (ii) was probabilistic sensitivity analysis (PSA) using 5,000 Monte Carlo iterations (seed 20260707), following ISPOR-SMDM good research practice.^28,35^ PSA input distributions were: Gamma on costs (standard error 20% of mean); Beta on utilities (method-of-moments from reported 95% CIs or 10% standard errors); Lognormal on hazard ratios (parameters from reported 95% CIs); Beta(k+1, n-k+1) posteriors on adverse-event rates from PROpel event counts^7^; and truncated Normal on adverse-event disutilities (20% relative standard error). Layer (iii) was distributional sensitivity across lognormal, GG and log-logistic parametrisations on the same Guyot pseudo-IPD (§3.5). Layer (iv) was structural comparison of PSM and Markov results (§3.2). Layer (v) was the methodological sensitivity analyses in §4. These were pseudo-IPD reconstruction (§4.1), PD-clipping frequency (§4.2), a conservative HR_OS = HR_PFS bound (§4.3), a PD-state cost multiplier stress test (§4.4), and a time-on-treatment reparametrisation decoupling drug cost from PFS occupancy (§4.5; multiplier τ scales the incremental drug-cost accrual, τ = 1.0 base case). They further comprised a one-way utility sensitivity (§4.6), an external-anchored OS extrapolation cap (§4.7), a budget-impact one-way tornado (§4.8) and a BRCAm control-curve scale-parameter (σ) one-way sensitivity (§4.9). We added analyses §4.5-§4.9 to bound, respectively, the treatment-duration, utility-alignment, long-tail-extrapolation, budget-assumption and control-curve scale-parameter uncertainties that we identified as the principal interpretive caveats of the analysis.

**Figure 2.**
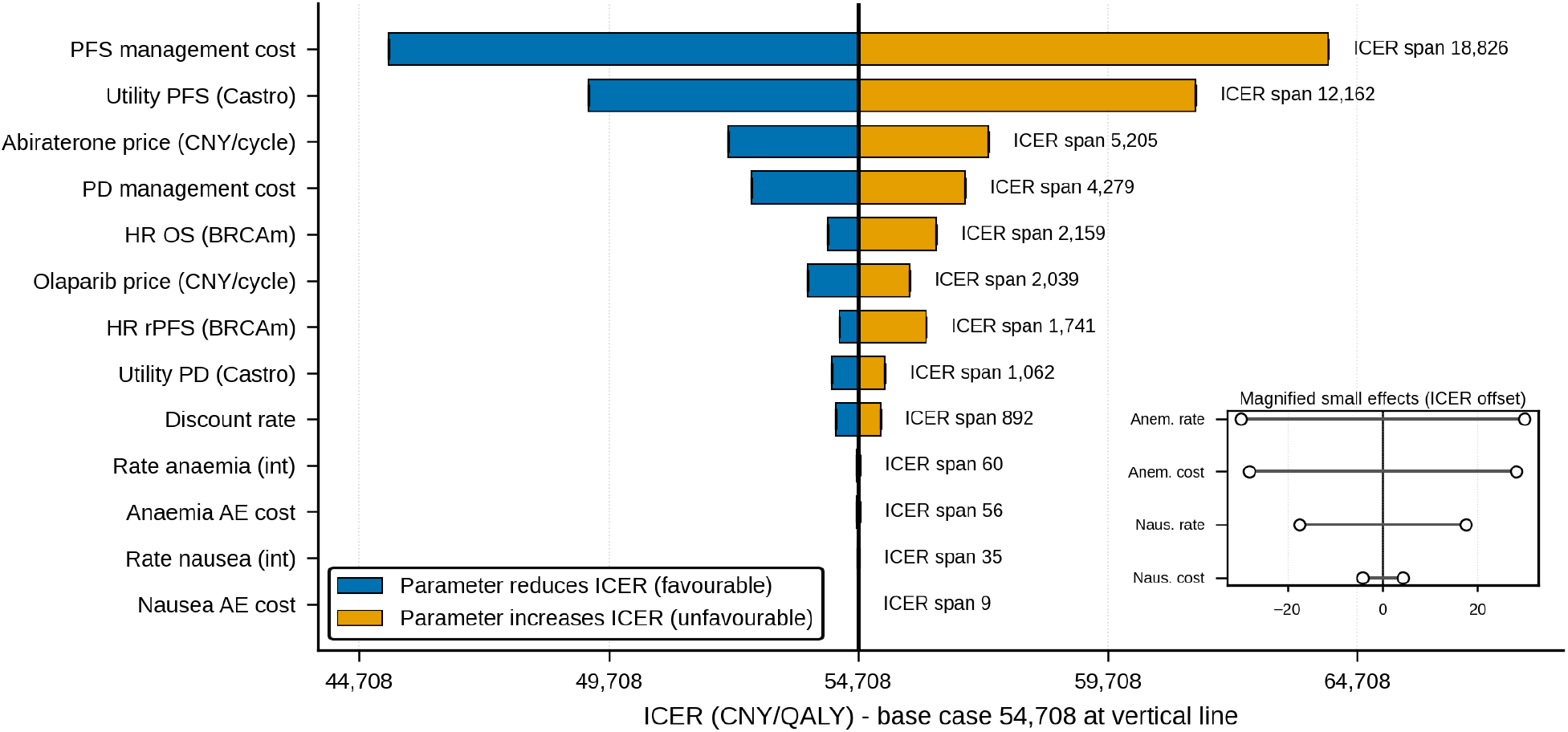
One-way deterministic sensitivity tornado analysis of the BRCAm × Castro base case, ranking 13 model parameters by absolute ICER impact when each is varied to its 95% confidence bound or ±20% around the point estimate while all others are held at base case. Bars are ordered top-down by absolute Δ ICER; the vertical line marks the base-case ICER of CNY 54,708/QALY, and the WTP threshold of CNY 287,247/QALY is off-scale to the right. PFS management cost is the single largest driver (Δ 18,826), followed by Castro PFS utility (Δ 12,162) and abiraterone price (Δ 5,205); the OS hazard ratio and olaparib price are minor drivers. The driver ranking is robust to the choice of subgroup survival inputs: the top two drivers (PFS management cost, PFS utility) are preserved when the ITT OS HR is used for the BRCAm subgroup, while abiraterone price and PD management cost exchange ranks.

We ran a supplementary PSA path (5,000 iterations, seed 42) to test BRCAm control-curve uncertainty and the effect of replacing proportional-hazards scaling with a non-PH structure. Path A (“PH-scaling”) jointly sampled BRCAm control-arm (μ, σ) parameters from asymptotic MLE bootstrap standard errors, BRCAm hazard ratios from lognormal draws and adverse-event rates from Beta(k, n−k) posteriors, retaining the PH scaling S_int(t) = S_ctrl(t)^HR. Path B (“AFT non-PH”) replaced PH scaling with an AFT intervention curve μ_int = μ_ctrl + log(shift), where we anchored the shift ratio to the base-case AFT-implied BRCAm median ratio (1/HR_OS = 1/0.29 = 3.448, giving 79.3 months) with a 15% coefficient of variation. Because the sparse BRCAm curves do not support a reliable direct paired PH diagnostic, activation of Path B used the reconstructed ITT OS log-cumulative-hazard slope ratio as a prespecified structural warning signal: we set the flag above 1.15, and the observed value was 1.185 (§3.1).

The value of information analysis computed per-patient expected value of perfect information (EVPI) as EVPI(WTP) = E[max(0, NMB)] − max(0, E[NMB]) across the PSA iterations,^31–32^ and per-patient expected value of partial perfect information (EVPPI) by nonparametric random-forest regression (Strong, Oakley and Brennan 2014^33^; scikit-learn RandomForestRegressor, 200 trees, min_samples_leaf 20, seed 20260707). EVPPI was computed across six parameter subsets: hazard ratios, utilities, drug costs, PFS management cost, post-progression cost, and adverse-event parameters. Because EVPI was zero at the CNY 287,247/QALY WTP threshold for all four scenarios (§3.4), we reported EVPPI at the peak-EVPI WTP per scenario. Because peak-EVPI WTP has no policy relevance in this decision, we present the EVPPI decomposition as an illustrative attribution of residual parameter uncertainty rather than as actionable evidence for research prioritisation. We report EVPPI subset shares as a range across the random-forest, out-of-bag and generalised-additive-model metamodels rather than as single point estimates. For metamodel robustness, we also computed each EVPPI subset under random-forest out-of-bag (OOB) predictions and under an independent generalised additive model (LinearGAM, pygam v0.12, 10 spline basis functions per feature), with subset-share sensitivity reported in Supplementary Table S1. We computed population EVPI over a 10-year horizon by discounting per-patient EVPI at 5% and multiplying by the annual incident eligible population.

The 5-year budget impact analysis followed a Chinese healthcare payer perspective. We based annual incidence on about 134,000 new prostate cancer cases in 2022^2–3^ with 1% growth. The BIA cascade multiplied incidence by 30% de novo metastatic proportion, 60% progression to castration resistance, 60% eligibility for first-line combination therapy, and 10% BRCA1/2 mutation prevalence.^7–8^ We assumed BRCA testing capacity would ramp from 30% to 70% over 2025-2029. Because the 10% prevalence is a Western tumour-based figure whereas Wei et al. report 5.6% germline BRCA1/2 prevalence in a large unselected Chinese cohort,^5–6^ we report a prevalence sensitivity analysis (Table 6 note and §5.4): substituting 5.6% for 10% scales the 5-year budget impact proportionally to CNY 13.46 million (low), 26.08 million (base) and 42.30 million (high uptake). The BRCA1/2 prevalence enters only the population/budget-impact layer and not the per-patient cost-utility model, so the base-case ICERs (CNY 54,708-68,000/QALY for BRCAm) are prevalence-independent and unchanged under either the 5.6% or the 10% assumption; prevalence affects only the size of the eligible cohort and therefore the absolute budget impact. We modelled three olaparib-abiraterone uptake trajectories: low (10-30%), base (20-50%) and high (40-70%). We derived annual per-patient costs from the PSM lifetime run and compared cumulative 5-year budget impact with the counterfactual scenario of all eligible patients receiving abiraterone alone. Model implementation used Python 3.11 with NumPy, SciPy, pandas and scikit-learn.

### 2.8 Data and code availability

We will share, upon publication, the reconstructed pseudo-individual patient data, deterministic base-case results, the full probabilistic sensitivity analysis output (5,000 iterations, seed 20260707), the EVPI/EVPPI results at the CNY 287,247/ QALY WTP threshold (including the RF/OOB/GAM metamodel comparison), the parametric-distribution robustness matrix, the Markov transition matrices, the budget-impact results, and the full Python source code that regenerates all base-case ICERs. Original PROpel individual-patient-level data are not publicly available. Table 1 provides all input parameters and their sources.

## 3. Results

Section 3 reports the base-case analysis and its default sensitivity envelope (deterministic one-way, probabilistic, structural PSM-vs-Markov, distributional, budget impact, downstream Pluvicto sequencing). Section 4 then reports four methodological sensitivity analyses tied to specific evidence-based assumptions of the model rather than to parameter uncertainty: Guyot pseudo-IPD reconstruction (§4.1), per-scenario PD-clipping frequency (§4.2), the conservative HR bound (§4.3) and the PD-cost multiplier stress test (§4.4). Readers looking for a headline may focus on §3.1 and §3.4; readers interested in the sensitivity of the base case to the underlying assumptions should read §4 alongside the corresponding parts of §5.

### 3.1 Base-case cost-utility

Table 2 reports the base-case results, which should be read together with the four methodological sensitivity analyses in §4 (survival-curve construction and its bootstrap, PD-clipping frequency, the conservative HR_OS = HR_PFS bound, and the PD-state cost stress test), because these qualify the headline estimates. In particular, the HR-bound analysis in §4.3 shows that the ITT ICERs rise by 78-119% under a conservative equal-hazard assumption, while the BRCAm ICERs rise by only 2-3%. The BRCAm conclusion is therefore far more stable than the ITT one. For the BRCAm subgroup we constructed the OS control curve with a median-anchored, maturity-constrained lognormal at the published control-arm median of 23.0 months (§2.5), because the BRCA1/2-confirmed subgroup (n=85) is too small for reliable Guyot digitisation; the ITT analysis used Guyot pseudo-IPD from the PROpel Number-at-Risk table (§2.5) re-fit under lognormal survival distributions in flexsurv v2.3.

In the BRCAm base case with Castro et al. utilities, olaparib plus abiraterone yielded 5.91 discounted QALYs at a total discounted cost of CNY 348,585, compared with 2.36 QALYs at CNY 154,372 for abiraterone alone. Incremental cost was CNY 194,214, incremental QALY 3.55, and the ICER was CNY 54,708/QALY (approximately 19% of the WTP threshold of CNY 287,247/QALY). Applying Xu 2022 utilities to the same median-anchored parametrisation reduced the incremental QALY gain to 2.86 and raised the ICER to CNY 68,000/QALY (23.7% of WTP). In the ITT scenario with Castro utilities, incremental cost was CNY 21,674 and incremental QALY 0.539, giving an ICER of CNY 40,198/QALY; the ITT × Xu ICER was CNY 43,721/QALY. All four base cases were cost-effective at the WTP threshold.

### 3.2 Structural comparison: PSM versus Markov

Table 3 reports the PSM and the co-primary two-transition Markov ICERs for the four scenarios. We ran both structures on the same control-arm parametrisation as the PSM (BRCAm median-anchored OS control median 23.0 months; ITT Guyot lognormal OS control median exp(3.5555) = 35.01 months), so the comparison isolates the effect of the structural specification from any parametric shape change. The two-transition Markov (detailed below) calibrated both the PFS→PD and PD→Death constant hazards so that each arm reproduced BOTH its median rPFS and its median OS, with a 3% per-year PFS-state background mortality (Chinese life-table for men aged 65-75) applied uniformly across arms and identical one-off (cycle 0) adverse-event costs and disutilities. Markov ICERs were 50-77% higher than the PSM equivalents in BRCAm and 100-140% higher in ITT: BRCAm × Castro CNY 81,878/QALY versus the PSM 54,708 (+50%) and BRCAm × Xu CNY 120,558 versus 68,000 (+77%); ITT × Castro CNY 80,479 versus 40,198 (+100%) and ITT × Xu CNY 104,927 versus 43,721 (+140%). The two-transition Markov is thus the most conservative of the three structural specifications examined. A single-transition Markov (which derives the PFS→PD transition from the PH-scaled PFS curve and calibrates only PD→Death to the median OS) reproduces the ITT results at CNY 55,860/QALY (Castro) and 66,991 (Xu). For the BRCAm intervention arm, however, it is undefined. The prespecified BRCAm intervention OS calibration target (79.3 months, obtained by the naive median-ratio mapping 23.0/0.29 and not an observed or PH-scaled median) lies far above the corresponding rPFS calibration target (34.8 months). No non-negative single PD→Death hazard can therefore reproduce the OS target from the PH-scaled PFS curve (the calibration hits a zero-hazard boundary). We thus do not report the single-transition structure for BRCAm and instead use the two-transition calibration as the co-primary Markov specification. The maximum ICER across all three structures and all four scenarios is CNY 120,558/QALY (BRCAm × Xu, two-transition Markov), 42.0% of the WTP threshold, so the cost-effectiveness conclusion is invariant to structural specification.

To probe whether the constant PD→Death hazard was itself driving the PSM-vs-Markov gap, we re-ran the single-transition Markov cohort under two alternative PD→Death hazard specifications. The first was a Weibull hazard h(t) = κλ(λt)^(κ-1) with fixed shape κ=1.5 (increasing hazard, consistent with NICE DSU TSD 21 recommendations for post-progression mortality^45^). The second was a Gompertz hazard h(t) = B·exp(γt) with fixed γ=0.02 per month (24% annualised rise, reflecting the age-related mortality trajectory in the mCRPC background). We report this shape test for the ITT population only, where the single-transition structure is well defined. It is not defined for the BRCAm subgroup, whose prespecified intervention OS calibration target (79.3 months) lies far above its corresponding rPFS calibration target (34.8 months), so that no single-transition PD→Death hazard reproduces the OS target (see above). For BRCAm the shape-of-hazard question is instead subsumed by the co-primary two-transition Markov already reported. Under Weibull PD→Death the ITT ICERs were CNY 58,681 (Castro) and 71,702 (Xu); under Gompertz they were CNY 58,861 and 72,008; against the constant-hazard single-transition baseline of CNY 55,860 and 66,991 this is a 4-7% shift. All six ITT single-transition configurations (Castro and Xu × three hazard specifications) remained below the WTP threshold, with the maximum ICER of CNY 72,008 (ITT × Xu, Gompertz) at 25.1% of WTP. The PSM-vs-Markov gap in ITT is therefore driven primarily by the structural constraint of an exponential PD-state exit in the Markov engine rather than by the specific functional form of the PD→Death hazard.

A further methodological question is whether a partial-calibration Markov — one that calibrates the PFS→PD transition to the median rPFS in addition to calibrating PD→Death to the median OS — would narrow the PSM-vs-Markov gap, and whether the two structures should be presented as co-primary. We therefore implemented a two-transition calibration in which we solved both the PFS→PD and PD→Death constant hazards simultaneously (by one-dimensional root-finding) so that each arm reproduces BOTH its median rPFS and its median OS exactly. In contrast to the single-transition calibration, which derives PFS→PD from the PH-scaled PFS curve and calibrates only PD→Death, this specification hits all median rPFS and OS targets. It produces no boundary solutions in any of the four arms. The achieved Markov calibration medians (model targets rather than observed arm medians) are: BRCAm control rPFS 8.0 / OS 23.0 months; BRCAm intervention rPFS 34.8 / OS 79.3; ITT control rPFS 19.8 / OS 35.0; ITT intervention rPFS 30.0 / OS 43.2. At the true BRCAm OS hazard ratio (0.29), the two-transition Markov is the most conservative of the three structures in both populations. For BRCAm the two-transition Markov produces the HIGHEST ICER of the three structures — CNY 81,878/QALY for BRCAm × Castro (28.5% of WTP) and 120,558 for BRCAm × Xu (42.0%) — above the PSM (54,708 and 68,000); the single-transition Markov is undefined for BRCAm. For ITT the two-transition Markov is likewise the highest, at CNY 80,479/QALY for ITT × Castro (28.0%) and 104,927 for ITT × Xu (36.5%), above both the PSM (40,198 and 43,721) and the single-transition Markov (55,860 and 66,991). The mechanism, verified by state-occupancy decomposition, is the same in both populations but is far more pronounced for BRCAm. In the two-transition Markov the intervention arm’s PD→Death hazard is calibrated to its median OS. For the BRCAm scenario the prespecified intervention OS calibration target (79.3 months, from the naive median-ratio mapping 23.0/0.29) sits about 45 months ABOVE the corresponding rPFS calibration target (34.8 months). The calibrated intervention PD→Death hazard is therefore low (0.0192/month), and intervention post-progression occupancy is LARGE (2.46 discounted person-years versus 1.28 in the control arm). The intervention arm accordingly accrues a large post-progression drug and management cost. Incremental cost rises to CNY 210,046 (versus CNY 194,214 in the PSM) while the incremental QALY gain (2.57 Castro) is slightly compressed relative to the PSM (3.55), so the two-transition Markov ICER rises above the PSM rather than falling below it. For ITT the corresponding Markov intervention OS calibration target (43.2 months) sits about 13 months above its rPFS calibration target (30.0 months), so the same mechanism operates more weakly. This is a structural finding driven by the survival inputs and the calibration targets, not by any engine artefact. We present the structural estimates as a co-primary range rather than privileging a single point: across all structures and all four scenarios every estimate remains cost-effective, and the maximum is CNY 120,558/ QALY (BRCAm × Xu, two-transition Markov), 42.0% of the WTP threshold. The cost-effectiveness conclusion is therefore invariant to structural specification and to Markov calibration depth. Figure 7 shows the structural estimates and their relationship to the 1× and 3× GDP willingness-to-pay thresholds across all four scenarios.

### 3.3 Deterministic one-way sensitivity

Figure 2 shows the one-way sensitivity tornado for the BRCAm × Castro base case. The ICER was most sensitive to PFS management cost (Δ ICER CNY 18,826), followed by PFS utility (Δ 12,162), abiraterone price (Δ 5,205), post-progression (PD) management cost (Δ 4,279), the BRCAm OS hazard ratio (Δ 2,159) and olaparib price (Δ 2,039), in that order. The ±20% range on any single parameter did not push the ICER above CNY 65,000/QALY, well below the CNY 287,247/QALY threshold.

### 3.4 Probabilistic sensitivity, cost-effectiveness acceptability and value of information

Two quantities in this section carry a policy-relevant interpretation that is not obvious from their names. The expected value of perfect information (EVPI) is the maximum amount a decision-maker would pay for a study that eliminated all parameter uncertainty; an EVPI of zero at a given WTP threshold means further research is not expected to change the reimbursement decision at that threshold, whatever it uncovered. The expected value of partial perfect information (EVPPI) is the same quantity restricted to a subset of parameters and identifies where any residual research value would concentrate.

Table 4 summarises the PSA (5,000 iterations, seed 20260707), and Figure 4 (cost-effectiveness planes) and Figure 3 (CEAC and EVPI curves) display it. The ICER of mean costs over mean QALYs was CNY 56,914/QALY for BRCAm × Castro (95% uncertainty interval CNY 37,420-81,808) and CNY 71,554/QALY for BRCAm × Xu (44,747-111,424); the corresponding PSA median ICERs were CNY 56,408 and 70,887/QALY. The ITT-scenario mean ICERs were driven strongly negative by 19.4% (Castro) and 19.9% (Xu) probability of dominance (positive incremental QALYs with negative incremental cost), so the ICER of means (CNY 40,813 for ITT × Castro; CNY 44,536 for ITT × Xu) and the median ICERs (CNY 44,993 and 47,626) are more representative summaries. The probability of cost-effectiveness at the WTP threshold of CNY 287,247/QALY was 100.0% for both BRCAm scenarios and 99.9% for both ITT scenarios by the net-monetary-benefit criterion. The Monte Carlo standard error at n=5,000 is about ±0.04 percentage points for a 99.9% probability, so the ITT values are consistent with a true probability well above 99% but not identifiable as exactly 100% from 5,000 draws. At the more restrictive 1× GDP threshold of CNY 95,749/QALY, the probability of cost-effectiveness was 99.7% for BRCAm × Castro, 90.4% for BRCAm × Xu, 98.4% for ITT × Castro and 87.0% for ITT × Xu (consistent with the CEAC^46^ in Figure 3 and the Table 4 net-monetary-benefit column). To confirm that these secondary-threshold probabilities are not sensitive to the single PSA seed, we repeated the PSA across five independent seeds (20260707-20260711, 5,000 iterations each). The across-seed range of the 1× GDP probability of cost-effectiveness was tight in every scenario: 99.6-99.8% for BRCAm × Castro, 90.2-91.0% for BRCAm × Xu, 98.2-98.4% for ITT × Castro and 86.7-87.4% for ITT × Xu, and median ICERs varied by less than 1% across seeds (for example BRCAm × Castro 55,929-56,408/QALY). The 86.7-87.4% multi-seed range for ITT × Xu confirms that the secondary-threshold probabilities are stable across seeds.

**Figure 3.**
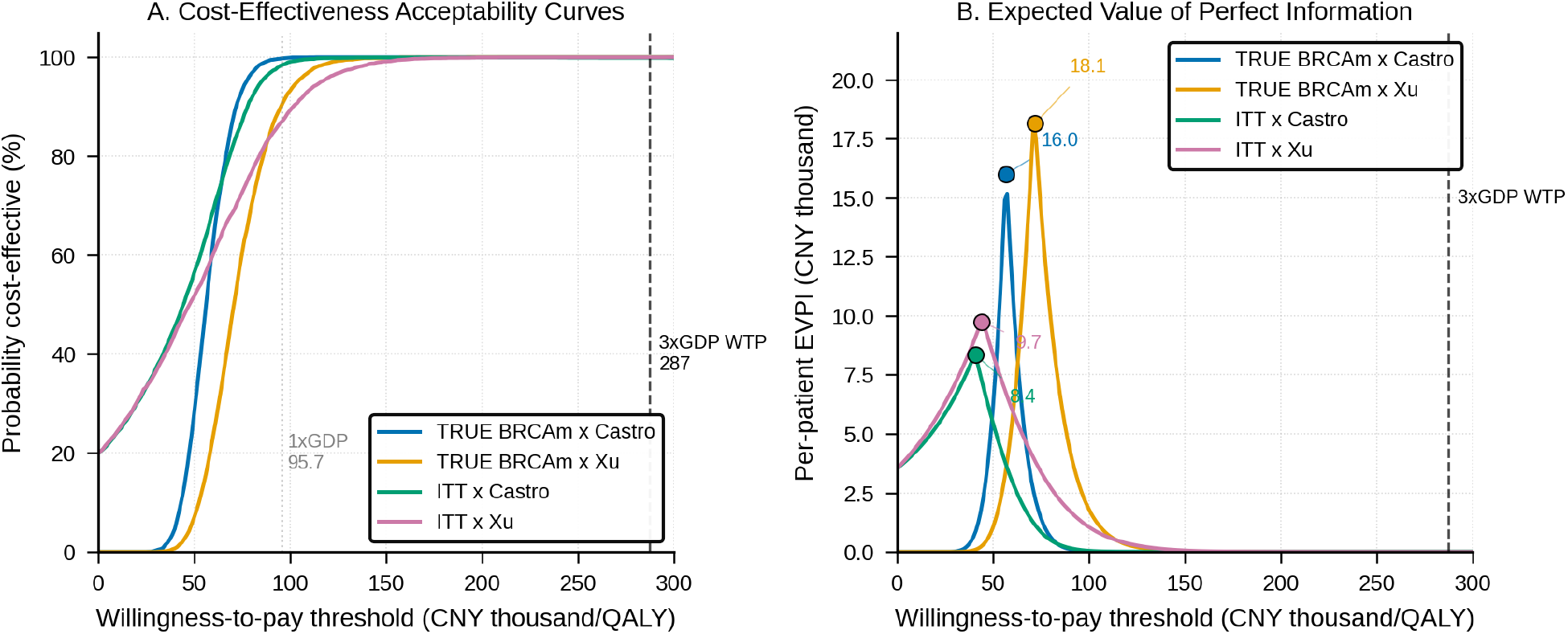
Cost-effectiveness acceptability curves for all four scenarios (left panel) and per-patient expected value of perfect information curves as a function of willingness-to-pay (right panel), showing the current Chinese WTP threshold of CNY 287,247/QALY. Peak EVPI is annotated for each scenario (BRCAm × Castro CNY 16,002 at WTP 57,000; BRCAm × Xu CNY 18,139 at WTP 72,000; ITT × Castro CNY 8,351 at WTP 41,000; ITT × Xu CNY 9,748 at WTP 44,000). All four scenarios reach ≥99% cost-effective probability at the 3× GDP WTP threshold and per-patient EVPI is zero at that threshold. The BRCAm curves were regenerated from the PSA; the ITT curves are carried forward unchanged (ITT invariant).

**Figure 4.**
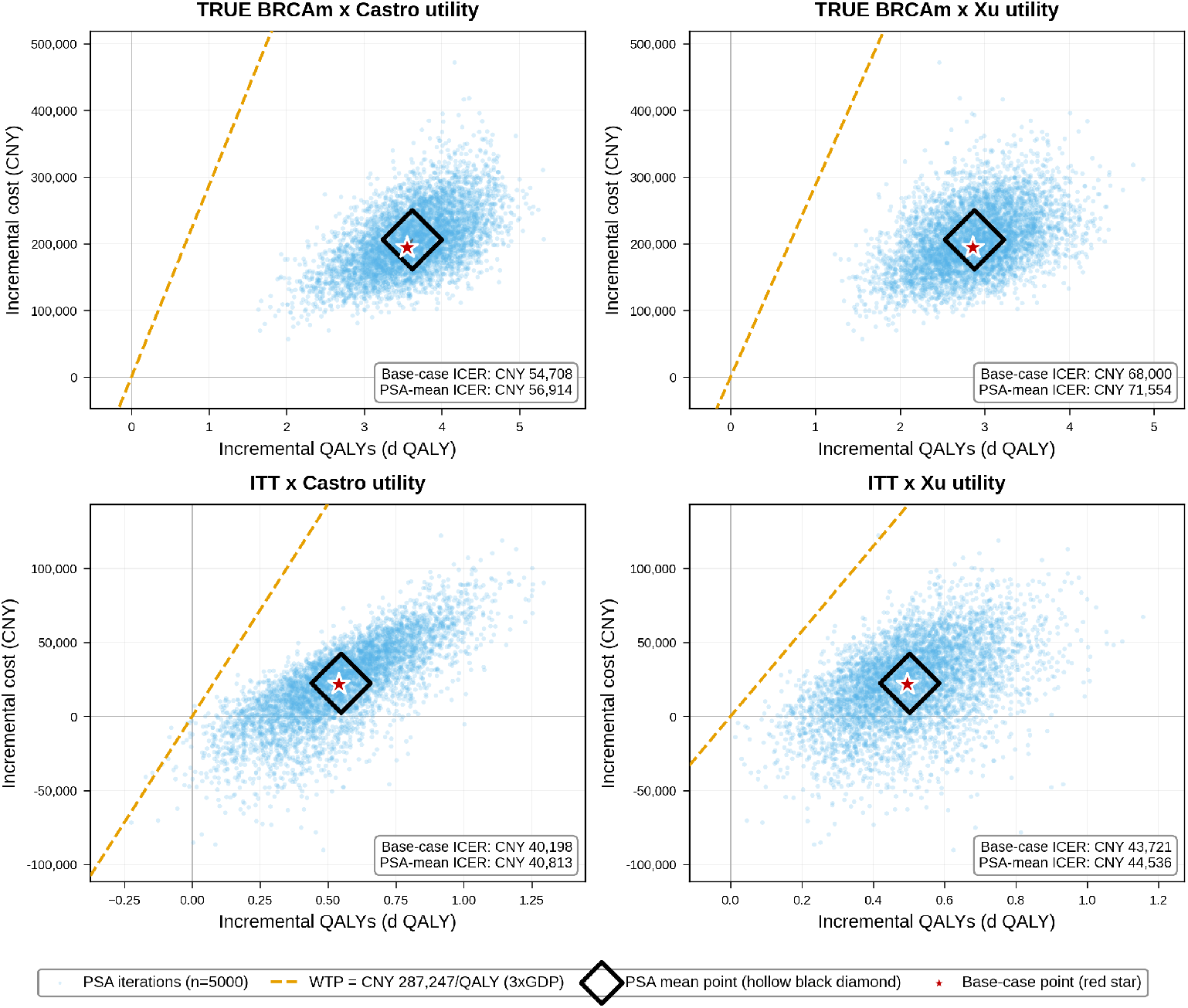
Cost-effectiveness plane scatter plots for all four scenarios (BRCAm × Castro, BRCAm × Xu, ITT × Castro, ITT × Xu), each showing 5,000 PSA iterations (light blue cloud) on the incremental-QALY (x-axis) versus incremental-cost (y-axis) plane at seed 20260707. **Red stars (★)** mark the deterministic base-case ICER for each scenario; **solid black diamonds (◆) mark the PSA mean-based ICER — that is, the ratio of the mean incremental cost to the mean incremental QALY across the 5,000 Monte Carlo iterations (ICER of means)** (not an error indicator). This ratio-of-means summary is reported in preference to the mean of the per-iteration ratios, which is undefined-in-the-limit and numerically unstable when incremental QALYs approach zero. Orange dashed lines depict the WTP threshold slope of CNY 287,247/QALY (3× 2024 China GDP per capita). Legend entries in each panel report both anchors: PSA mean-based ICERs (56,914; 71,554; 40,813; 44,536) and base-case ICERs (54,708; 68,000; 40,198; 43,721). The WTP slope is steep and lies at the upper-left corner of the BRCAm panels because the BRCAm base ICER (∼55-72k) is far below the WTP threshold.

Per-patient expected value of perfect information (EVPI) at the WTP threshold was effectively zero in every scenario, because the CEAC crossed 99.9% well before the threshold (Figure 3, right panel). Peak per-patient EVPI occurred at the WTP value adjacent to each scenario’s base-case ICER: CNY 16,002 for BRCAm × Castro at WTP CNY 57,000/QALY; CNY 18,139 for BRCAm × Xu at WTP CNY 72,000/ QALY; CNY 8,351 for ITT × Castro at WTP CNY 41,000/QALY; CNY 9,748 for ITT × Xu at WTP CNY 44,000/QALY (Table 5). Nonparametric EVPPI at the peak-EVPI WTP identified PFS management cost as the dominant contributor to residual decision uncertainty in the BRCAm scenarios (CNY 13,330, 83% of BRCAm × Castro peak EVPI; CNY 13,012, 72% of BRCAm × Xu). Utilities contributed 30% and 46%, hazard ratios 34% and 38%, drug costs 27% and 23%, and PD cost 23% and 20% respectively. In the ITT scenarios the ranking reversed: hazard ratios dominated (CNY 6,922, 83% of ITT × Castro peak EVPI; CNY 8,419, 86% of ITT × Xu), with PFS management cost contributing 40% and 35%, PD cost 36% and 32%, and drug costs 20% and 18%. The dominance of PFS management cost in the BRCAm setting is mechanistic. At HR_PFS 0.23 applied to a control-arm median rPFS of 8 months, the intervention arm accrues about 7.35 discounted years in PFS versus 2.42 years in control (an incremental 4.93 discounted years, or about 64 discounted cycles). Any per-cycle uncertainty in PFS management cost is therefore amplified by this large multiplier. In ITT, where the incremental QALY gain is much smaller (0.539 vs 3.55), the survival-modelling parameters instead drive uncertainty. Adverse-event parameters contributed 12-15% of scenario EVPI in every case, consistent with the negligible ICER sensitivity to nausea rate framing noted in §2.6.

The subset-share ranking was stable across the three EVPPI metamodels of §2.6 (RF in-sample, RF out-of-bag [OOB], LinearGAM). In 20 of 24 subset-scenario combinations, the OOB share differed from the in-sample RF share by less than 1 percentage point (maximum divergence 2.53 pp); the four exceptions were all the adverse-event subset, where the OOB share was about 2.3 percentage points lower than the RF share. The dominant subset was identical across the three metamodels in every scenario (PFS management cost in BRCAm; hazard ratios in ITT), and the top-two ranking was preserved throughout (Supplementary Table S1). The GAM produced systematically lower subset shares (subset sums 172-193% vs 203-219% for RF), consistent with GAM’s additive-only structure missing the parameter interactions that RF captures, but the ranking was unchanged. The subset sum above 100% is a structural property of the non-orthogonal parameter partitions in the Strong-Oakley-Brennan framework^33^ and reflects joint information across subsets (i.e., pairs of parameters can jointly contain information beyond either individually) rather than evidence of metamodel overfitting; each subset EVPPI is individually bounded above by total scenario EVPI.

Over a 10-year discounted horizon, the eligible BRCAm-identified population was about 7,394 patients and the ITT-eligible population about 126,164 patients. Peak population EVPI reached CNY 134 million for BRCAm × Xu at WTP CNY 72,000/ QALY and CNY 1,230 million for ITT × Xu at WTP CNY 44,000/QALY; at the CNY 287,247/QALY threshold, population EVPI was zero in every scenario.

### 3.5 Distributional and non-PH sensitivity

We assessed distributional robustness differently for the two populations, reflecting the difference in survival-data maturity described in §2.5. For the ITT population, we fitted alternative parametric distributions (lognormal, generalised gamma (GG), log-logistic) to the Guyot pseudo-IPD in flexsurv v2.3, following NICE DSU TSD 14 recommendations for multi-distribution reporting^29^; we evaluated the fitted distributions over the 15-year horizon, and Figure 6 and Supplementary Table S2 summarise the resulting four-specification ICER envelope. For the BRCAm subgroup, the multi-distribution matrix is not applicable. As detailed in §2.5, the BRCAm placebo arm (n=38, 25 OS events) is too sparse to fit or discriminate among competing parametric tails. The BRCAm base case is therefore the single median-anchored lognormal curve, and we quantify its parametric uncertainty probabilistically via the PSA rather than through a distributional matrix. Supplementary Table S2 therefore reports the four ITT distributions in full (Panel B) but, for BRCAm, reports only the median-anchored base case alongside its PSA-derived uncertainty interval.

**Figure 5.**
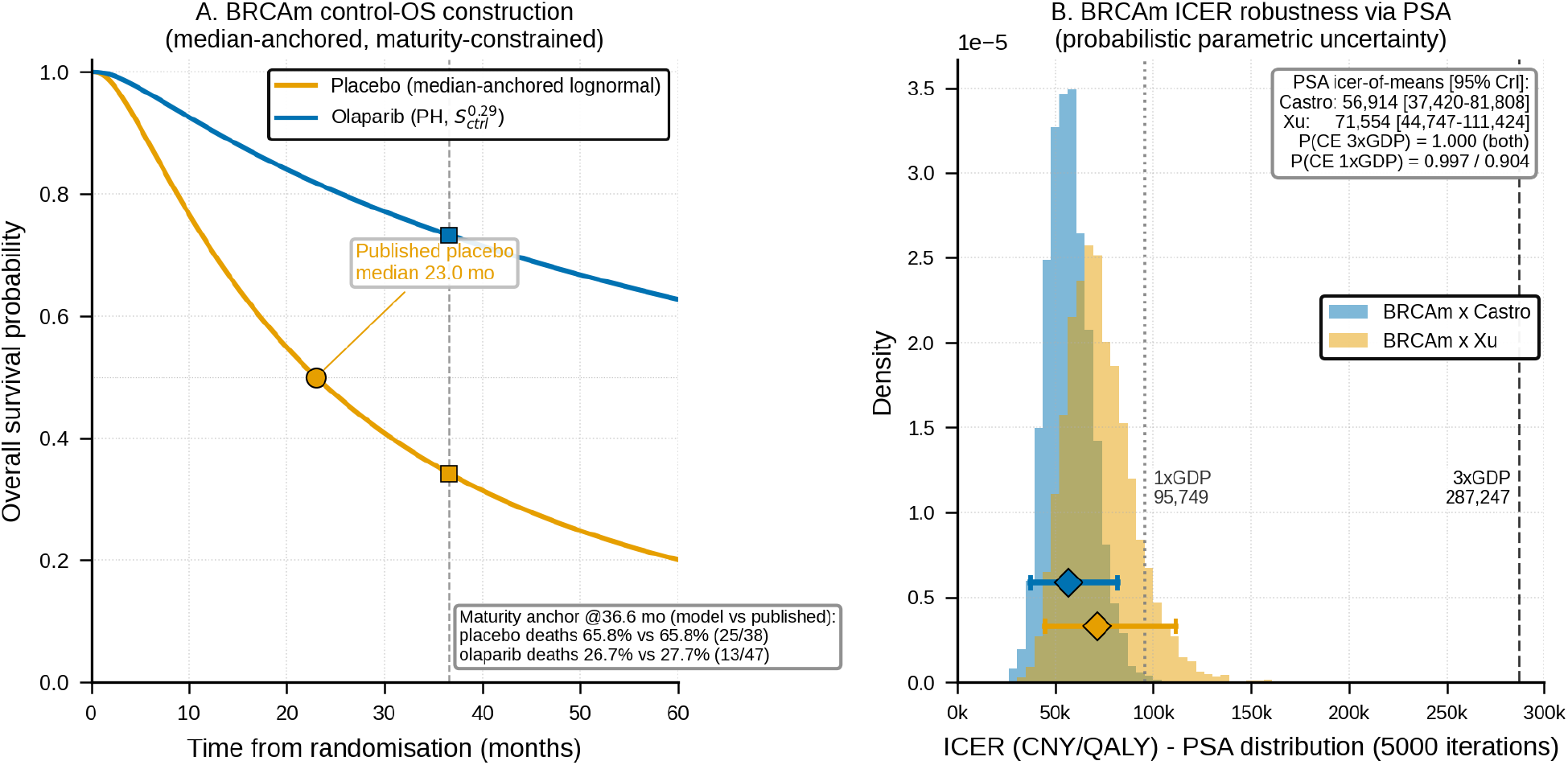
BRCAm subgroup control-arm OS construction and parametric-uncertainty robustness (FUSION approach, §2.5). Because the BRCAm placebo arm (n=38, 25 OS events) is too sparse for reliable Guyot digitisation or multi-distribution parametric selection (NICE DSU TSD 14), the BRCAm base case uses a single median-anchored, maturity-constrained lognormal control curve rather than a set of Guyot-fitted alternatives. Panel A: the median-anchored control-arm OS curve, a lognormal fixed to the published BRCAm control median OS of 23.0 months (meanlog μ = ln 23.0 = 3.1355 log-months) with the scale parameter σ = 1.1422 solved so the survivor function reproduces the observed control mortality at the data cut-off (S(36.6 months) = 1 − 25/38 = 0.342); the maturity-anchor points (published median 23.0 months and the 36.6-month cut-off mortality) are marked. Under proportional-hazards scaling with the true BRCAm OS HR of 0.29 the modelled intervention OS median is ∼105 months (23.0/0.29 = 79.3 months on the naive AFT anchor; 105 months as the median of the PH-scaled curve), consistent with the published intervention median being not reached. Panel B: PSA-based ICER robustness for the BRCAm subgroup — the distribution of the BRCAm × Castro and BRCAm × Xu ICERs across the 5,000-iteration PSA (seed 20260707), which quantifies BRCAm parametric uncertainty probabilistically in place of the invalid Guyot multi-distribution matrix. The 95% credible intervals (Castro CNY 37,420–81,808; Xu CNY 44,747–111,424) lie entirely below the WTP threshold.

**Figure 6.**
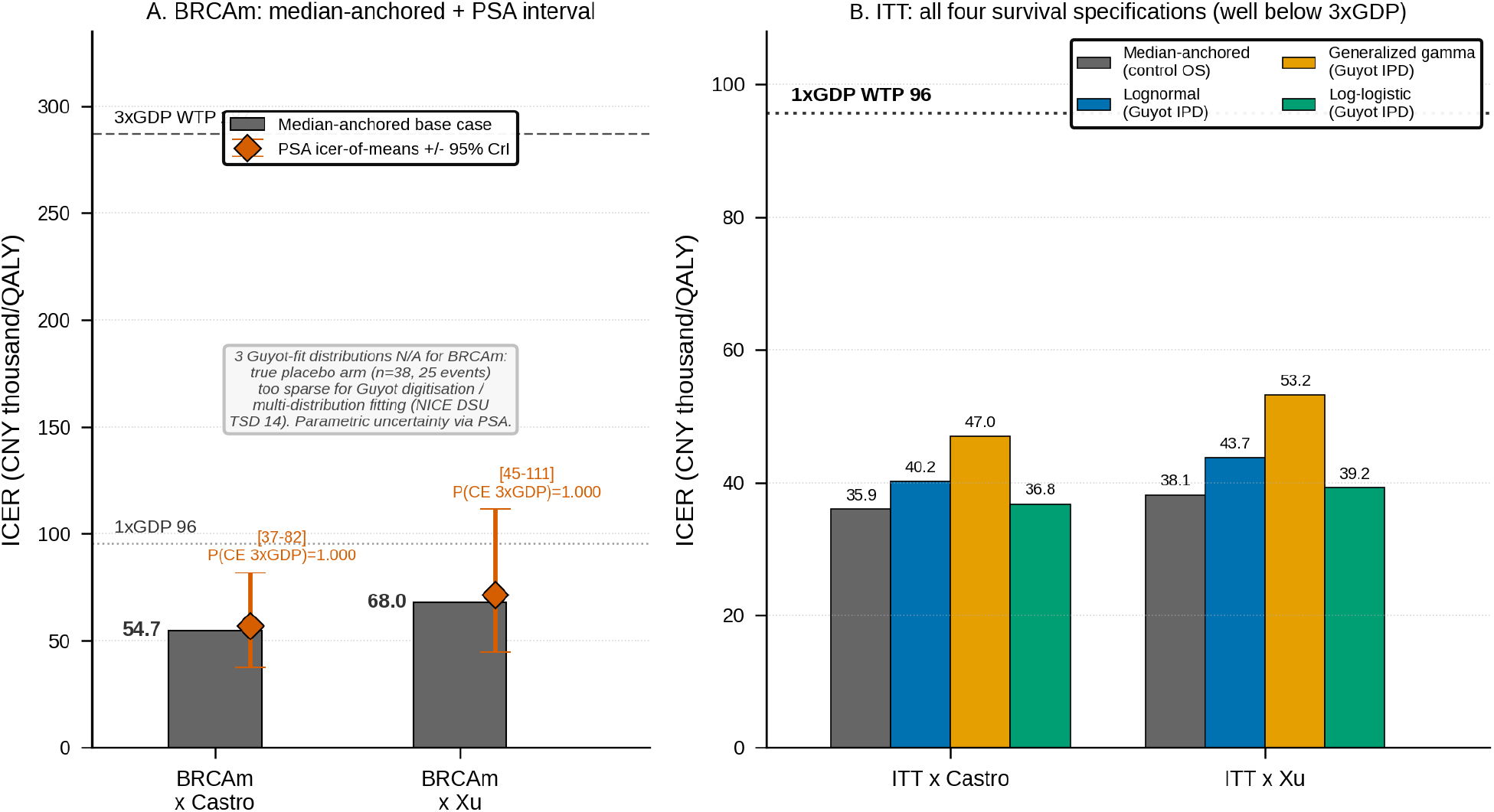
Parametric-distribution robustness of the ICER (CNY thousand/QALY). Panel A (BRCAm subgroup): the median-anchored lognormal base case is shown as bars (Castro CNY 54,708; Xu CNY 67,999) with the PSA icer-of-means and its 95% credible interval overlaid as a diamond with error bars (Castro CNY 37,420–81,808, P(CE at 3× GDP) = 1.000; Xu CNY 44,747–111,424, P(CE at 3× GDP) = 1.000). The three Guyot-fitted parametric alternatives (lognormal/generalised gamma/log-logistic) are not applicable to BRCAm because the BRCAm placebo arm (n=38, 25 OS events) is too sparse for Guyot digitisation or multi-distribution discrimination (NICE DSU TSD 14), so BRCAm parametric uncertainty is quantified probabilistically via the PSA instead (§2.5, §3.5); reference lines at 1× GDP (CNY 95,749) and 3× GDP (CNY 287,247). Panel B (ITT population): ICER under all four OS survival specifications — median-anchored (control OS), Guyot lognormal base case, Guyot generalised gamma, and Guyot log-logistic — for the Castro and Xu utility configurations (Castro CNY 35,922 / 40,198 / 46,955 / 36,765; Xu CNY 38,101 / 43,721 / 53,209 / 39,187), because the ITT Guyot pseudo-IPD (n=397 placebo, 205 events) support valid multi-distribution fitting; all specifications lie well below the 1× GDP reference line (CNY 95,749) and hence far below 3× GDP.

**Figure 7.**
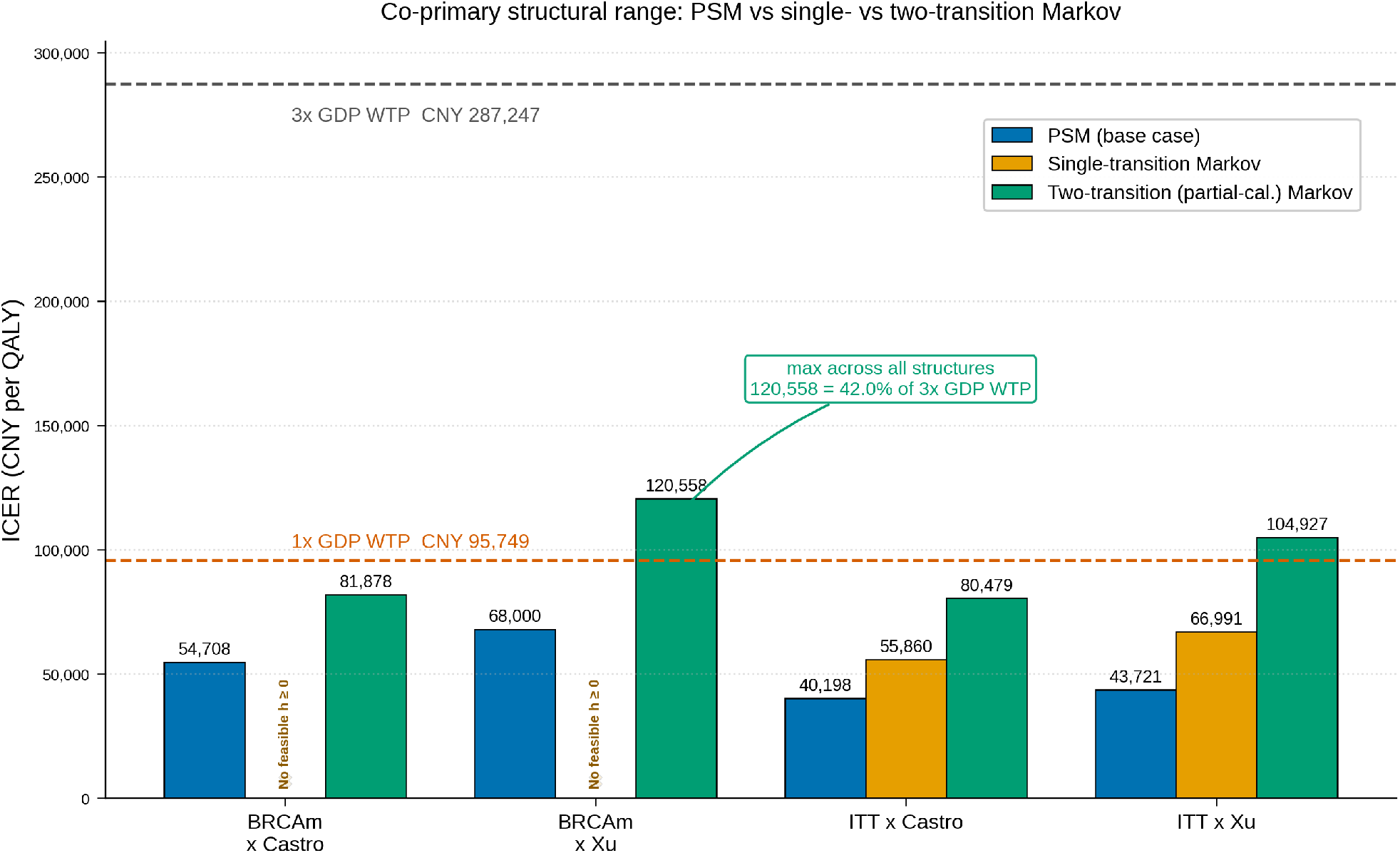
Co-primary structural range of the incremental cost-effectiveness ratio across the four scenarios (BRCAm × Castro, BRCAm × Xu, ITT × Castro, ITT × Xu) under three model structures: the partitioned survival model (PSM) base case, the single-transition Markov (PD→Death calibrated to median OS only) and the two-transition Markov (both PFS→PD and PD→Death calibrated to median rPFS and median OS; §3.2). Under the true BRCAm final OS hazard ratio (0.29) the two-transition Markov is the HIGHEST of the three structures for BOTH populations (Markov/PSM ratio 1.50–1.77 for BRCAm, 2.00–2.40 for ITT): because the prespecified Markov BRCAm intervention OS calibration target (79.3 months = 23.0/0.29, a naive median-ratio anchor rather than an observed or PH-scaled median) far exceeds the corresponding rPFS target (34.8 months), the intervention arm accrues a long, costly post-progression occupancy, so the transition-based structure raises rather than lowers the BRCAm ICER — the reverse of the ordering obtained when the ITT hazard ratios are applied to the BRCAm subgroup. The single-transition Markov reproduces the ITT results but is undefined (zero-hazard boundary) for the BRCAm intervention arm, because no non-negative single PD→Death hazard can reproduce an OS median of 79.3 months from the PH-scaled rPFS curve; this boundary is annotated in the figure. Dashed lines mark the 1× GDP (CNY 95,749) and 3× GDP (CNY 287,247) willingness-to-pay thresholds. The maximum ICER across all structures and scenarios is CNY 120,558/QALY (BRCAm × Xu, two-transition Markov), 42.0% of the 3× GDP threshold, so the cost-effectiveness conclusion is invariant to structural specification.

For the BRCAm subgroup, we propagated parametric uncertainty through the main PSA and through a supplementary PSA (Supplementary Table S2 Panel A) rather than through a distributional matrix. The main PSA (5,000 iterations, seed 20260707) yielded an ICER of mean costs over mean QALYs of CNY 56,914/ QALY for BRCAm × Castro (95% credible interval 37,420-81,808) and CNY 71,554/QALY for BRCAm × Xu (95% CrI 44,747-111,424), with cost-effectiveness probability at the WTP threshold of 100% in both cases. These intervals quantify parametric uncertainty within the prespecified median-anchored construction, but they do not eliminate residual distributional model-form uncertainty. The BRCAm base case therefore remains cost-effective across the full range of probabilistically sampled control-curve shapes.

The ITT distributional envelope was wider, and we report it in full. Under Castro utilities, ITT ICERs ranged from CNY 36,765/QALY (log-logistic, −8.5% vs lognormal) through CNY 40,198 (lognormal, base case) to CNY 46,955/QALY (GG, +16.8%); under Xu utilities they ranged from CNY 39,187 (log-logistic) through CNY 43,721 (lognormal) to CNY 53,209/QALY (GG, +21.7% vs lognormal). Across all four ITT survival specifications (median-anchored, lognormal, GG, log-logistic) the ITT ICERs spanned CNY 35,922-46,955/QALY under Castro and 38,101-53,209/QALY under Xu, i.e. the upper bound of each range exceeded its lower bound by 30.7-39.7%. The larger ITT event sample (n=205 events over the observed follow-up) resolves distributional differences more sharply than the BRCAm subgroup, yet even the widest ITT distributional ICER (CNY 53,209/QALY under ITT × Xu × GG) was 18.5% of the WTP threshold.

For the BRCAm subgroup a supplementary PSA path (5,000 iterations, seed 42; Supplementary Table S2 Panel A) additionally replaced PH scaling with an AFT intervention curve μ_int = μ_ctrl + log(shift), where the shift equals 1/HR_OS = 1/0.29, giving an AFT intervention-arm OS median of 79.3 months; the PH curve retains its approximately 105.2-month median. On the median-anchored BRCAm control curve, the AFT-path mean ICER was CNY 56,172/QALY (95% CrI 41,541-69,151) versus the PH-path mean of CNY 56,461/QALY (95% CrI 40,498-73,185), a difference of only 0.5%; both paths had cost-effectiveness probability ≥99.8% at 1× GDP and 100% at ≥1.5× GDP. The near-equality of the resulting mean ICERs is therefore an empirical robustness result rather than a consequence of equal survival medians: despite the 105.2-versus 79.3-month structural difference, the BRCAm cost-effectiveness conclusion is unchanged.

### 3.6 Budget impact

Table 6 summarises the 5-year budget impact. Under the base uptake trajectory (olaparib plus abiraterone reaching 20% of eligible patients in 2025 and 50% by 2029), cumulative 5-year budget impact was CNY 46.57 million, versus counterfactual all-comer control costs of CNY 330.1 million. The low-uptake trajectory (10-30%) gave CNY 24.03 million and the high-uptake trajectory (40-70%) gave CNY 75.53 million. These estimates reflect the true BRCAm OS HR (0.29): under the strong modelled OS benefit a substantial fraction of intervention patients remain alive and on therapy within the undiscounted 5-year window, so the per-patient incremental cost over that window is material (an undiscounted 5-year incremental cost of about CNY 95,000 per patient; see §5.2 for the mechanism). Drug acquisition during the extended PFS period drove the budget impact primarily; post-progression management costs contribute less because the strong OS benefit keeps patients in PFS longer within the 5-year horizon. The BIA is sensitive to the assumed BRCA testing capacity ramp (30-70%) and to the 60% eligibility fraction, both flagged in §5.4. Because the same first-line BRCAm cohort will later be eligible for Pluvicto on progression, §3.7 reports the downstream sequencing budget impact under the current pricing environment as a separate expenditure category.

To test whether the deterministic BIA understates uncertainty by varying only the testing ramp and uptake, we first rebuilt the BIA with an exact per-patient-year cohort-tracking engine. This engine reproduces the corrected deterministic estimates to within 0.05 million (low CNY 24.03 million, base 46.57 million, high 75.53 million). We then propagated all six cascade parameters jointly by Monte Carlo (10,000 simulations, seed 20260717). We drew each cascade parameter from a PERT distribution anchored on the published point value: probability of metastatic disease at diagnosis PERT(0.25, 0.30, 0.40), progression to mCRPC PERT(0.50, 0.60, 0.70), first-line combination eligibility PERT(0.45, 0.60, 0.75), and BRCA1/2 prevalence PERT(0.056, 0.10, 0.12). The prevalence range spans the Chinese germline lower bound of 5.6%^5–6^ to the Western tumour-based figure of 10-12%^7–8^. We scaled the testing ramp by a Uniform(0.70, 1.15) multiplier (capping the annual testing rate at 0.80). Under joint cascade uncertainty the 5-year cumulative budget impact had a mean of CNY 21.88 million (95% interval 13.16 to 33.36) under low uptake, 42.42 million (25.51 to 64.67) under base uptake and 68.79 million (41.37 to 104.88) under high uptake. After pooling the 10,000 draws from all three uptake scenarios, the pooled cross-scenario 95% interval was CNY 14.9 to 94.0 million (full pooled range 9.1 to 151.3 million). This percentile interval is not the union of the three scenario-specific 95% intervals. The probabilistic means sit slightly below the deterministic points because the BRCA-prevalence distribution is left-skewed toward its 0.10 mode. All scenarios remain within a magnitude routinely absorbed in provincial reimbursement, confirming that the budget-impact conclusion is robust to joint cascade uncertainty and not an artefact of varying only two inputs. Figure 8 shows the probabilistic means, 95% intervals, p10–p90 ranges and deterministic anchors for the three uptake trajectories, together with this pooled cross-scenario 95% interval. We treat the cascade parameters as independent, a standard simplification.

The testing-capacity ramp itself (30% in 2025 to 70% by 2029) is a policy scenario rather than an empirical forecast, and no single published Chinese prostate-cancer BRCA/HRR testing-uptake rate exists to calibrate it against. The direction and starting level of the ramp are nonetheless supported by the Chinese evidence base. A nationwide survey and external quality assessment of 32 third-party next-generation-sequencing laboratories in China documented a low and highly heterogeneous testing landscape, with an overall external-quality-assessment pass rate of about 60% and testing concentrated in small and medium laboratories with under five years of operation and no national HRR-testing guideline at the time.^47^ A real-world north-China cohort reported HRR mutation detection in only 3% of tumour tissue and 3.9% of blood samples and explicitly characterised uptake as relatively low, calling for better pre-test candidate selection.^48^ And a recent review of prostate cancer in China concluded that real-world uptake and access to BRCA/ HRR testing lag Western benchmarks.^49^ International benchmarks are consistent with a sub-50% starting point and room to grow. A cross-sectional physician survey spanning the USA, EU5, Japan and China found that only about half of mCRPC patients are recommended for genetic testing, with persistent patient- and physician-level barriers,^50^ A real-world analysis reported that fewer than half of mCRPC patients were tested for HRR mutations.^51^ We therefore read the 30%-to-70% ramp as a transparent policy assumption bracketing a documented low, heterogeneous baseline with substantial room for expansion, and the probabilistic analysis above quantifies the BIA’s robustness to it (testing-ramp scale varied by Uniform(0.70, 1.15)).

### 3.7 Downstream Pluvicto sequencing budget impact

This subsection is an order-of-magnitude sizing exercise rather than a formal cost-effectiveness analysis of Pluvicto: we model no incremental effectiveness, utility or comparator survival for the radioligand pathway, and the scenario grid is intended only to place the downstream expenditure envelope on the same scale as the first-line decision. Across an 18-scenario grid combining BRCA testing coverage (30%/ 70%), Pluvicto uptake (5%/10%/20%) and unit price (CNY 80,000/100,000/150,000 per dose; six doses per course), the annual eligible-for-Pluvicto flow ranged from 8 to 76 patients per year and total annual budget impact ranged from CNY 3.8 million to CNY 68.4 million. Cumulative five-year expenditure in the steady state ranged from CNY 19.2 million to CNY 342.0 million, an 18-fold spread. The worst-corner annual expenditure (CNY 68.4 M) is comparable to the first-line olaparib-abiraterone base-uptake five-year cumulative BIA (CNY 46.57 M) and is dominated by per-dose price and uptake rather than by BRCA testing coverage. Readers should read these figures as a resource-planning sizing exercise, not as evidence on the value of Pluvicto itself; Supplementary Table S3 gives full scenario detail.

## 4. Methodological sensitivity analyses

Four pre-specified methodological sensitivity analyses complement the base-case results.

### 4.1 Survival-curve construction: ITT Guyot reconstruction and BRCAm median anchoring

The two populations require different survival-curve construction methods because they differ markedly in the maturity of the published Kaplan-Meier data. For the ITT population, to avoid the circularity of fitting alternative parametric distributions to pseudo-IPD simulated from a base-case lognormal MLE, we reconstructed the ITT control-arm Kaplan-Meier data with the Guyot algorithm^24^ from the published Number-at-Risk table.^7^ Reconstruction produced ITT-consistent pseudo-IPD with OS event counts within one event of the published totals (177 vs 176 OS-olaparib; 206 vs 205 OS-placebo), and the ITT rPFS reconstruction reproduced the published rPFS event counts of 246 (olaparib) vs 271 (placebo) at the DCO3 data cut-off (median follow-up about 36.6 months). Independent lognormal, GG and log-logistic re-fits in flexsurv v2.3 on the resulting pseudo-IPD serve as the ITT parametric distributions (§2.5; Supplementary Table S2 Panel B).

We cannot reconstruct the BRCA1/2-confirmed subgroup the same way: the published BRCAm placebo arm (n=38, 25 OS events)^9^ is far too sparse to digitise reliably or to discriminate among competing parametric tails. We therefore constructed the BRCAm OS control curve by median anchoring with a maturity constraint; the intervention arm applies the published BRCAm OS HR of 0.29 by proportional-hazards scaling. BRCAm rPFS is likewise median-anchored, with the published rPFS HR of 0.23 applied by PH scaling. This is because the BRCAm subgroup rPFS Kaplan-Meier curves are not published at Guyot-digitisable resolution^4,9,7^ (construction detailed in §2.5). All four base-case ICERs (CNY 54,708, 68,000, 40,198, 43,721/QALY) remained cost-effective at the WTP threshold.

Because the BRCAm control curve is median-anchored rather than digitised, a nonparametric bootstrap of a BRCAm pseudo-IPD is not available (there is no valid n=85 digitisation to resample). We instead propagate the finite-sample and parametric uncertainty that a bootstrap would represent probabilistically in the main PSA (Table 4). The PSA samples the BRCAm OS HR from its published 95% confidence interval (0.29, 0.14-0.56), the rPFS HR from (0.23, 0.12-0.43) and all cost, utility and adverse-event parameters from their priors, anchored to the correct BRCA1/2 population. Under this probabilistic uncertainty the BRCAm × Castro ICER had a 95% credible interval of CNY 37,420-81,808 (P(cost-effective at 3× GDP) = 1.000) and BRCAm × Xu 44,747-111,424 (1.000), confirming that the BRCAm conclusion is robust to control-curve and parameter uncertainty.

### 4.2 PD-clipping frequency across scenarios

The PSM cycle-loop uses the standard clipped-subtraction identity p_pd = max(p_os − p_pfs, 0), which zeroes out post-progression occupancy when the intervention arm’s PH-scaled S_OS(t) crosses below its S_PFS(t). Supplementary Table S2 Panel B reports the per-scenario crossing frequency for both arms (“Cross I/C” column) for the four ITT distributions, and the accompanying note reports it for the BRCAm median-anchored base case. Crossing was zero in both arms for ITT × lognormal (base case) and for ITT × median-anchored; 1 cycle for ITT × log-logistic; and 125 intervention / 92 control cycles for ITT × GG (driven by the shorter GG tail). In the BRCAm median-anchored base case, crossing occurred at 156 intervention / 130 control cycles (80% / 67% of the 195-cycle horizon), clipping about 86% of the would-be post-progression QALY mass. This is a material point for interpretation: PD-clipping is essentially a BRCAm-subgroup phenomenon and is absent (0 of 195 cycles) from the primary ITT base case, so the ITT result — which we assigned the primary decision-analytic weight in this analysis — does not depend on the clipped-subtraction identity at all. The clipping caveat therefore applies to the supporting BRCAm scenario, not to the headline ITT conclusion. BRCAm clipping is a consequence of the intervention arm’s long lognormal OS tail (intervention-arm OS median approximately 105.2 months under PH scaling at HR_OS 0.29) combined with the shorter rPFS tail, which under PH scaling produces S_int_PFS(t) > S_int_OS(t) at longer horizons where the residual population is small. Because BRCAm PD occupancy is clipped to zero across most horizon cycles, the BRCAm incremental QALY gain is driven almost entirely by the pre-progression (PFS) state, and post-progression utility assumptions have minimal impact on the BRCAm ICER. The complementary HR bound sensitivity in §4.3 quantifies the effect of setting HR_OS equal to HR_PFS; because the true BRCAm hazard ratios are already close (0.23 and 0.29), this bound moves the BRCAm ICER only marginally.

### 4.3 Hazard-ratio bound sensitivity

Because the BRCAm subgroup rPFS Kaplan-Meier curves are not published at Guyot-digitisable resolution^9,7^ and we constructed the BRCAm control-arm OS curve by median anchoring rather than digitisation (§2.5), we cannot perform a paired log-cumulative-hazard PH diagnostic on the BRCAm subgroup. The BRCAm base case therefore relies on the PH assumption via S_int(t) = S_ctrl(t)^HR, with the reported BRCAm HR_PFS = 0.23 and final HR_OS = 0.29.^9,7^ As a bound sensitivity, we recomputed all four base cases with HR_OS set equal to HR_PFS (BRCAm HR_OS = 0.23 instead of 0.29; ITT HR_OS = 0.66 instead of 0.81), corresponding to the more aggressive assumption that the intervention’s OS benefit is at least as steep as its rPFS benefit. The bound is stronger than the published HR_OS point estimates (BRCAm final 0.29, 95% CI 0.14-0.56; ITT 0.81, 95% CI 0.67-1.00)^9,7^ and we use it solely to quantify the ICER sensitivity to the direction of any residual PH departure; it is not a plausibility claim about the true OS effect. Two directional properties should be noted before reading the numbers. First, although the parameter change is optimistic about the intervention’s OS benefit, the resulting ICERs are higher (more conservative on the ICER scale) because incremental cost scales faster than incremental QALYs in this parameter region, which is why the abstract, introduction and policy sections describe the bound as a conservative HR_OS = HR_PFS bound. Second, the magnitude of the ICER shift under this bound is not uniform across scenarios: it is very small in BRCAm and large in ITT, reflecting how far apart the OS and rPFS hazard ratios are in each population. Under the bound, BRCAm × Castro ICER shifted from CNY 54,708 to CNY 55,854/QALY (+2.1%) and BRCAm × Xu from CNY 68,000 to CNY 69,905/QALY (+2.8%); the BRCAm result was highly stable. ITT × Castro shifted from CNY 40,198 to CNY 71,652/QALY (+78.2%) and ITT × Xu from CNY 43,721 to CNY 95,872/QALY (+119.3%); the ITT result was highly sensitive to the HR_OS assumption. All bound-sensitivity ICERs remained below the WTP threshold. The true PROpel hazard ratios now explain the contrast between subgroups. In BRCAm the OS and rPFS hazard ratios are already close (0.29 and 0.23), so forcing them equal barely changes the intervention-arm OS extrapolation and hence the ICER. In ITT, by contrast, the gap between HR_OS (0.81) and HR_PFS (0.66) is large, so the equal-hazard bound substantially steepens the ITT OS curve and raises the ITT ICER. Figure 9, Panel A shows the full propagation of the BRCAm OS-HR confidence interval.

### 4.4 PD-state cost multiplier stress test

Because the one-way tornado (§3.3) identified PD-state monthly cost as a material ICER driver (the fourth-ranked parameter in the BRCAm × Castro scenario and the largest among the post-progression management-cost inputs), we recomputed the four base cases across seven PD-cost multipliers {0.5×, 0.8×, 1.0×, 1.2×, 1.5×, 2.0×, 3.0×}. We did so by deterministic re-scoring of the same PSM cycle loop (no new PSA; all other parameters at base-case values). The 1.0× reference corresponds to the base-case PD-state management cost of CNY 5,831.53 per cycle (§2.6). This cost is itself built up from docetaxel chemotherapy applied to 50% of the post-progression cohort as a conservative assumption, plus continuing supportive care, rather than an arbitrary equal-to-PFS-cost assumption. The PFS-state management cost is CNY 2,595.75 per cycle, so the base case already assigns post-progression management a 2.25× higher per-cycle cost than the pre-progression state. This stress test brackets that value from 0.5× to 3.0× to ensure the base multiplier is justified rather than assumed. This is a deterministic point-estimate sensitivity rather than a probabilistic replication: we provide no PSA envelope, and the reported dominance cells at ITT × 2.0× and ITT × 3.0× reflect the deterministic point estimate under stressed PD cost rather than a probabilistic uncertainty band. Incremental QALYs are invariant across multipliers by construction (state occupancy and utilities do not depend on state cost); because incremental cost is likewise utility-independent, the incremental-cost value in each dominant cell is a single figure shared by both utility sources. The 1.0× row reproduces the base-case ICERs to the cent (CNY 54,708 / 68,000 / 40,198 / 43,721/QALY) as an internal reproducibility check.

All 28 ICERs remained below the CNY 287,247/QALY threshold. The maximum stressed BRCAm × Castro ICER at 0.5× was CNY 60,057/QALY (∼21% of WTP); the maximum stressed BRCAm × Xu at 0.5× was CNY 74,648/QALY (∼26% of WTP). The direction is monotonically decreasing in the multiplier: raising PD-state cost makes intervention relatively less costly than control because olaparib plus abiraterone extends PFS and shortens PD, so incremental cost falls as PD cost rises. The BRCAm slope is shallow: because the true BRCAm intervention arm spends most of the horizon in the pre-progression state (PH-scaled intervention-arm OS median approximately 105.2 months), post-progression cost changes have limited leverage, and even at a 3× over-estimation of PD-state cost the BRCAm scenarios remain cost-effective (CNY 33,313 Castro / 41,407 Xu, ∼12-14% of WTP). In the ITT scenarios, incremental cost crosses zero between the 1.5× and 2.0× multipliers, so the intervention becomes strictly dominant at multipliers ≥ 2.0× under both utility sources.

### 4.5 Time-on-treatment sensitivity

The base-case PSM accrues intervention drug cost over full pre-progression (PFS) occupancy, implicitly assuming treatment continues until radiographic progression. Because real-world discontinuation before progression (adverse events, patient preference, non-progression clinical events) shortens actual time on treatment, we ran a one-way sensitivity that decouples intervention drug-cost accrual from PFS. It applies a multiplier τ to the incremental drug component (τ = 1.0 base case; τ = 0.85 and 0.70 representing 15% and 30% average reductions in on-treatment time relative to PFS). Because τ < 1 lowers only intervention drug cost, the ICER improves monotonically as modelled treatment duration shortens: ITT × Castro fell from CNY 40,198/QALY (τ = 1.0) to CNY 28,844 (τ = 0.85) and CNY 17,490 (τ = 0.70); ITT × Xu behaved analogously. BRCAm × Castro fell from CNY 54,708 to CNY 51,020 and CNY 47,332; BRCAm × Xu from CNY 68,000 to CNY 63,416 and CNY 58,832. All values remained below 1× GDP. The base case (τ = 1.0) is therefore the conservative bound on this axis (Figure 9, Panel C; Supplementary Table S5): modelling real-world discontinuation would improve, not worsen, cost-effectiveness. The PROpel final safety analysis reported adverse-event-driven discontinuation of 13.8% (olaparib arm) vs 7.8% (placebo arm) at median follow-up 36.6 months,^52^ consistent with τ modestly below 1 in practice; the base case does not exploit this favourable direction.

### 4.6 One-way utility sensitivity

To test the dependence of the conclusion on the imperfectly aligned utility sources (§2.6), we varied each PFS and PD utility by ±0.05 around its source-specific central value, one at a time, holding all other inputs at base case. The resulting ICER ranges were CNY 36,546-44,661/QALY (ITT × Castro), CNY 39,435-49,052/QALY (ITT × Xu), CNY 51,153-58,794/QALY (BRCAm × Castro) and CNY 62,593-74,429/QALY (BRCAm × Xu). Every one-way utility perturbation left all four scenarios below the WTP threshold and preserved the ordering ITT < BRCAm, confirming that the cost-effectiveness conclusion and the relative ranking of the scenarios are robust to plausible utility mis-specification (Figure 9, Panel D; Supplementary Table S6). The two independent utility sources (Castro meta-analytic; Xu Chinese) already bracket the result, and this one-way analysis quantifies the local gradient within each source.

### 4.7 OS extrapolation cap (external-anchored survival truncation)

To confirm that the supporting BRCAm result is not an artefact of the long modelled OS tail (PH-scaled intervention-arm median of about 105 months; §5.3), we capped the intervention-arm OS median to progressively shorter values by substituting weaker OS hazard ratios. The caps were a mid cap at 60 months (HR_OS ≈ 0.43), the MAGNITUDE IPCW-adjusted BRCA1/2 median of 36.7 months (HR_OS 0.645), and the MAGNITUDE unadjusted median of 29.4 months (HR_OS 0.788). The BRCAm × Castro ICER was flat across this range: CNY 54,708 (105 months, base) → 53,808 (60 months) → 53,432 (36.7 months) → 53,328/QALY (29.4 months). BRCAm × Xu behaved analogously (CNY 68,000 → 65,908/QALY at the shortest cap). The near-invariance arises because truncating the OS tail removes both incremental cost and incremental QALYs in the low-survival-probability region roughly proportionally. This directly addresses the concern that the BRCAm cost-effectiveness conclusion depends on extrapolated long-term survival: it does not (Figure 9, Panel B; Supplementary Table S7).

**Figure 8.**
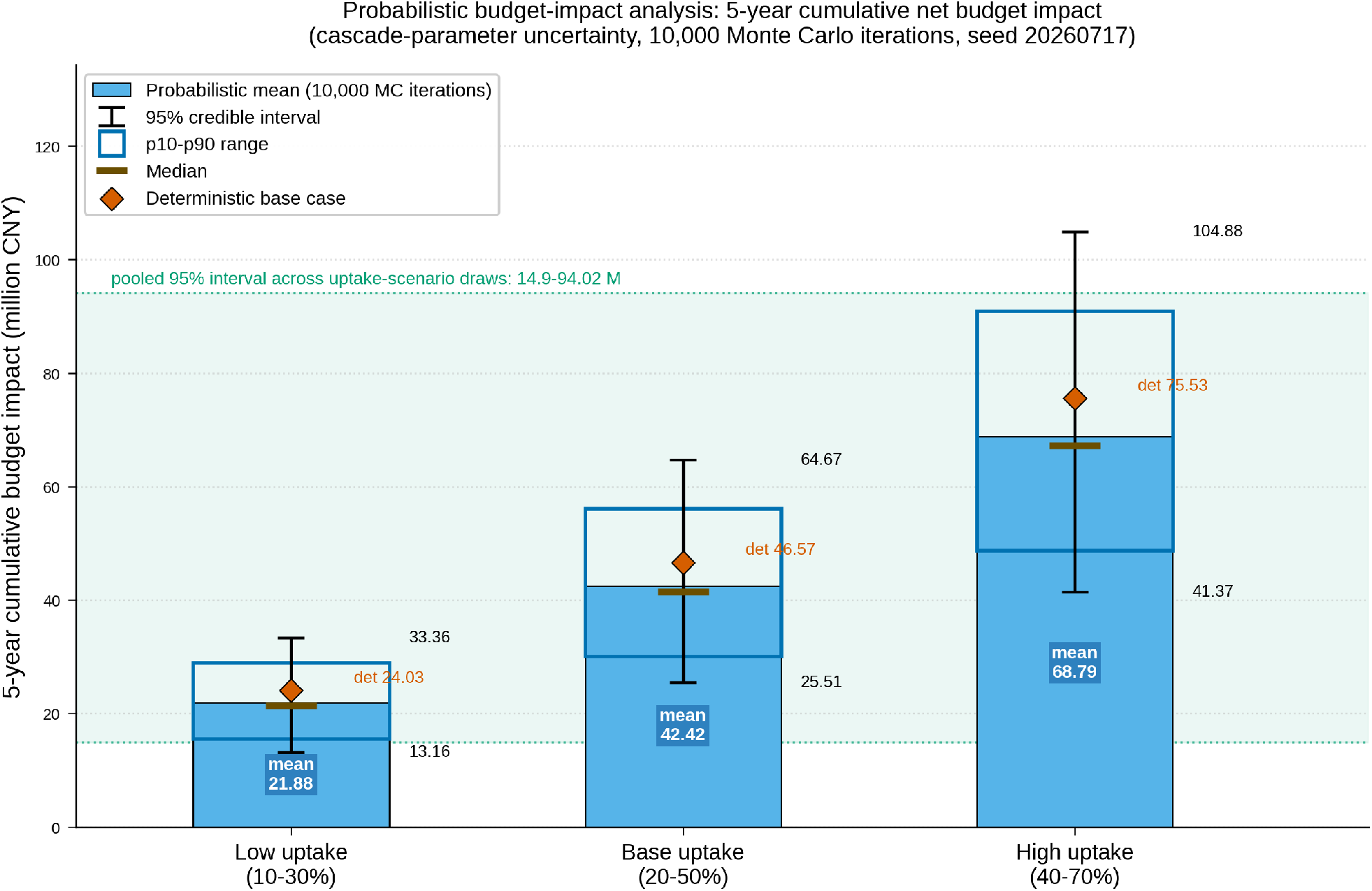
Probabilistic budget-impact analysis (§3.6). Five-year cumulative net budget impact under the three olaparib–abiraterone uptake trajectories (low 10–30%, base 20–50%, high 40–70%), with all six BIA cascade parameters propagated jointly by Monte Carlo (10,000 iterations, seed 20260717) on an exact per-patient-year cohort-tracking engine that reproduces the published deterministic BIA to within 0.1%. Blue bars are probabilistic means (CNY 21.88, 42.42 and 68.79 million); black whiskers are 95% credible intervals (13.16–33.36, 25.51–64.67 and 41.37–104.88 million); dark-blue box outlines are the p10–p90 range; white ticks are medians; orange diamonds are the deterministic base-case estimates (CNY 24.03, 46.57 and 75.53 million). The light-green band is the pooled 95% interval after combining the Monte Carlo draws from the three uptake scenarios (CNY 14.9–94.0 million); it is not the union of the scenario-specific 95% intervals (which would span CNY 13.16–104.88 million). These values are computed on the BRCAm-specific OS HR of 0.29 and are about five-fold larger than the estimate obtained when the ITT OS HR of 0.66 is applied to the BRCAm subgroup (base CNY 9.41 million), because the strong BRCAm-specific OS benefit keeps more intervention patients alive and on therapy within the undiscounted 5-year window.

**Figure 9.**
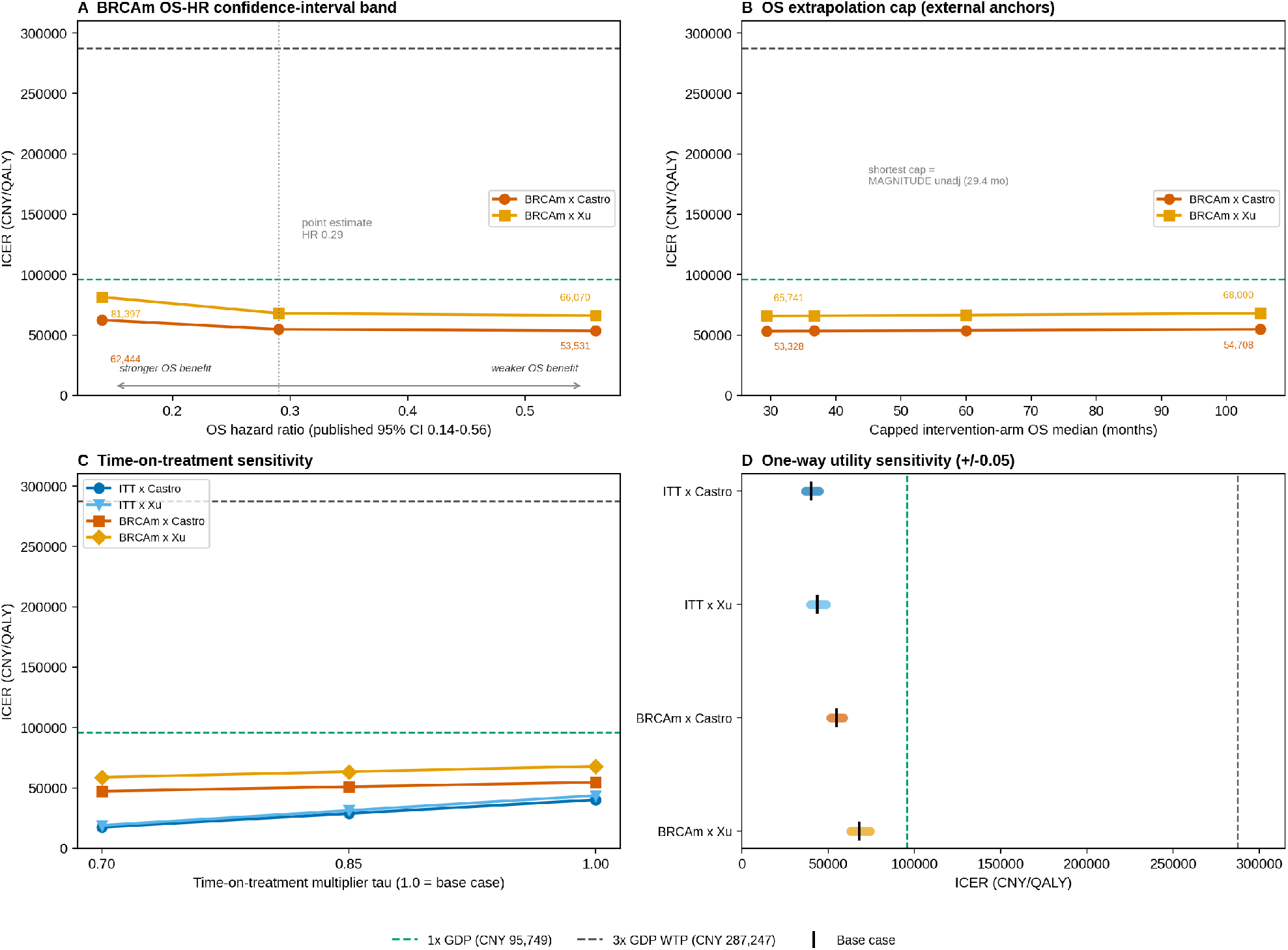
Bounding-robustness panel (§4.5–4.8). Deterministic ICERs under four bounding sensitivity analyses, all referenced to the 1× GDP line (CNY 95,749, teal dashed) and the 3× GDP willingness-to-pay threshold (CNY 287,247, grey dashed). **Panel A** — the supporting BRCAm ICER across the full published OS-HR 95% confidence interval (0.14–0.56): BRCAm × Castro CNY 62,444 (HR 0.14) to 53,531 (HR 0.56) with the point estimate 54,708 at HR 0.29; BRCAm × Xu analogously 81,397 to 66,070. **Panel B** — BRCAm ICER when the modelled intervention-arm OS median is capped from about 105 months down to 60, 36.7 (MAGNITUDE IPCW-adjusted) and 29.4 months (MAGNITUDE unadjusted) by substituting weaker OS hazard ratios; the near-flat profile (BRCAm × Castro 54,708→53,328) shows the result is not tail-driven. **Panel C** — time-on-treatment sensitivity: ITT × Castro falls from CNY 40,198 (τ = 1.0) to 17,490 (τ = 0.70) and BRCAm × Castro from 54,708 to 47,332 as modelled treatment duration shortens; the base case (τ = 1.0) is the conservative bound. **Panel D** — one-way ±0.05 utility ranges for all four scenarios, with base-case markers (black); every range and the ordering ITT < BRCAm sit below the WTP threshold. Every point in all four panels is below 3× GDP, and the primary ITT scenarios remain below 1× GDP throughout.

### 4.8 Budget-impact one-way sensitivity

Complementing the joint probabilistic cascade analysis (§3.6), a one-way tornado ranked the five cascade policy inputs by their impact on the 5-year base-uptake budget (CNY 46.57 million), varying each across its PERT/uniform support. BRCA1/2 prevalence was the dominant driver (5-year budget CNY 26.08-55.89 million across prevalence 0.056-0.12; swing CNY 29.8 million), followed by metastatic-disease probability and combination-eligibility fraction (each swing about CNY 23.3 million), the testing-ramp scale (swing about CNY 21.0 million) and progression-to-mCRPC probability (swing about CNY 15.5 million). All one-way excursions remained within a range routinely absorbed in provincial reimbursement. This makes explicit that the budget inputs are transparent policy parameters with disclosed ranges, and identifies BRCA prevalence — a measurable epidemiological quantity — as the most valuable target for Chinese real-world calibration (Figure 10; Supplementary Table S8).

**Figure 10.**
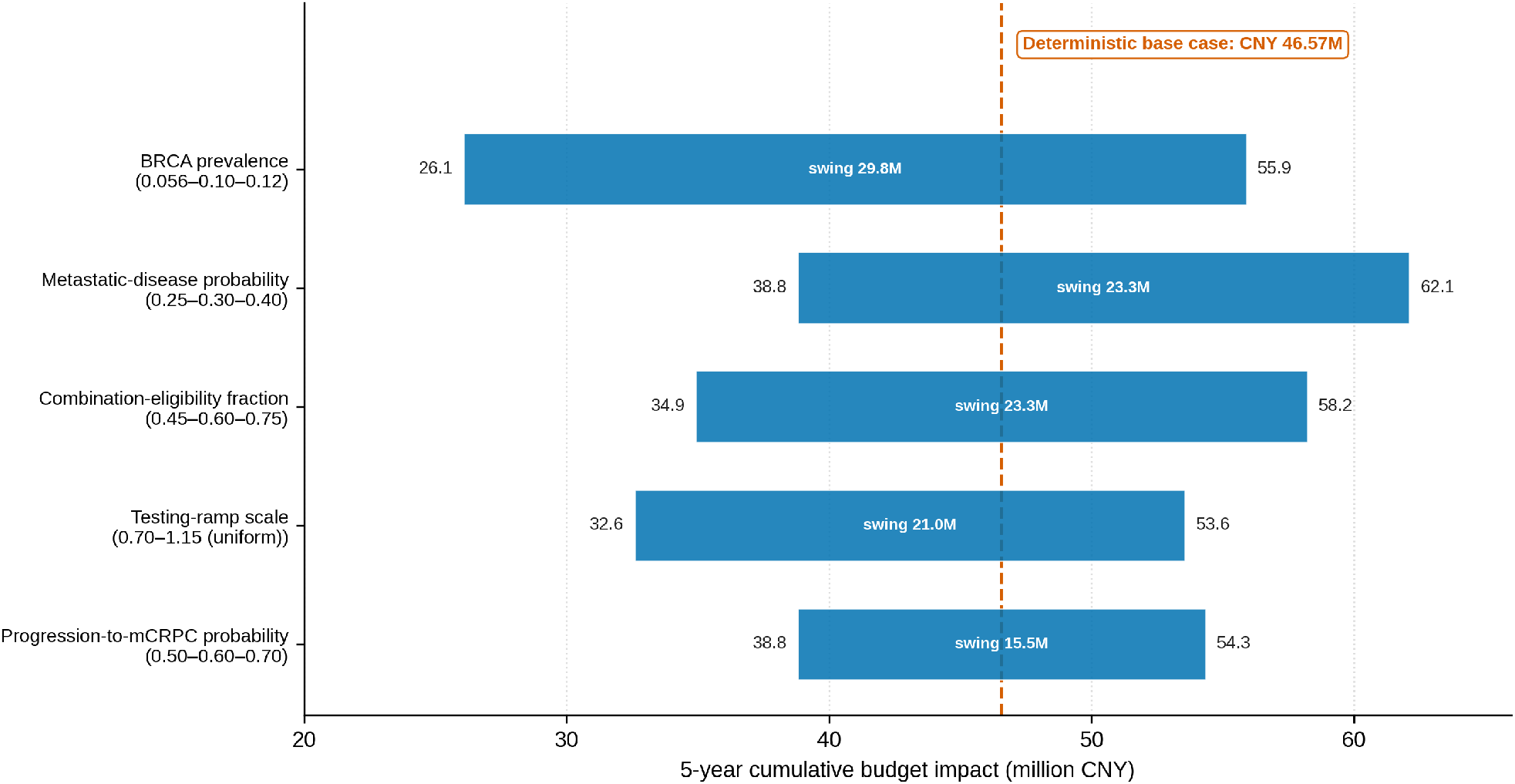
Budget-impact one-way tornado (§4.8). Each of the five cascade policy parameters was varied one at a time across its PERT/uniform support, holding all others at base case, and the resulting 5-year cumulative payer budget impact (million CNY) is shown as a horizontal bar spanning the low and high values; bars are ordered by swing magnitude (largest driver at top). The orange dashed line marks the deterministic base-case budget (CNY 46.57M). BRCA1/2 prevalence is the dominant budget driver (swing CNY 29.8M) — a measurable epidemiological quantity and the highest-value target for Chinese real-world calibration — followed by metastatic-disease probability and combination-eligibility fraction (each swing about CNY 23.3M), testing-ramp scale (CNY 21.0M) and progression-to-mCRPC probability (CNY 15.5M). This one-way tornado ranks drivers by magnitude and does not replace the joint probabilistic BIA (§3.6). Supporting values in Supplementary Table S8.

### 4.9 BRCAm control-curve scale-parameter (σ) one-way sensitivity

The BRCAm control-arm OS curve is the median-anchored, maturity-constrained lognormal constructed in §2.5, whose scale parameter σ = 1.1422 was solved from the published control-arm maturity rather than fitted to individual-patient data. Because σ is therefore a modeller-derived quantity rather than a directly estimated one, we propagate its uncertainty jointly in the PSA (§3.4) but additionally interrogated it here as an explicit one-way deterministic analysis, to match the per-parameter convention of §4.3–§4.6. Holding the published median (μ = ln 23.0) and all other inputs at base case, we varied σ by ±20%. The BRCAm × Castro ICER moved from the base CNY 54,708/QALY to CNY 54,943/QALY at σ − 20% (σ = 0.914) and CNY 53,968/QALY at σ + 20% (σ = 1.371). This is a total one-way range of CNY 53,968–54,943/QALY (at most +0.4% from base). BRCAm × Xu behaved analogously (CNY 66,784–68,388/QALY; at most +0.6%), with the maximum σ-excursion ICER reaching only 23.8% of the WTP threshold. As a data-anchored alternative to the arbitrary ±20% span, we re-solved σ so the control curve reproduces the upper Clopper-Pearson 95% bound of the observed control mortality (0.804, giving σ = 0.544). This changed the ICERs by less than 0.1% (BRCAm × Castro CNY 54,687; BRCAm × Xu CNY 67,965). The lower mortality bound (0.486) is structurally unreachable while holding the published 23.0-month median, because the 36.6-month cut-off lies beyond the median, so control survival at cut-off cannot exceed 0.5 for any σ. This property shows σ is in fact tightly identified from above once the published median is fixed. The near-invariance of the ICER to σ has the same mechanistic origin as σ’s low rank in the Figure 2 tornado. Because PD-state occupancy is clipped to zero in about 80% of horizon cycles for the BRCAm intervention arm (§4.2), the BRCAm incremental QALY is driven almost entirely by pre-progression time. The shape of the control-arm OS tail (the only thing σ controls) therefore has minimal leverage on the BRCAm ICER (Supplementary Table S9).

## 5. Discussion

### 5.1 Principal findings

At current post-VBP prices, first-line olaparib plus abiraterone is cost-effective for first-line mCRPC in China. We frame the intention-to-treat (ITT) population as the primary decision-analytic result because it rests on the larger PROpel evidence base (N=796; 205 control-arm OS events observed over follow-up), and we report the prespecified BRCA1/2-mutated (BRCAm) subgroup as a supporting precision-medicine analysis that aligns with the NMPA-approved indication, although a small subgroup (n=85) informs it. The ITT base-case ICER was CNY 40,198/QALY under meta-analytic utilities and CNY 43,721/QALY under Chinese utilities, both below 1× the 2024 Chinese per-capita GDP (CNY 95,749) and about 14-15% of the 3× GDP WTP threshold. The ITT result was robust across the four survival specifications (CNY 35,922-53,209/QALY), to a ±0.05 utility swing (CNY 36,546-49,052/QALY, §4.6), and to time-on-treatment reparametrisation (§4.5). The partitioned-survival PD-clipping concern does not affect the ITT result at all: the ITT intervention arm has zero cycle-crossings over the 15-year horizon (§4.2), so the ITT ICER does not rely on the clipped-subtraction identity.

In the supporting BRCAm subgroup the ICER was CNY 54,708/QALY (meta-analytic utilities) and CNY 68,000/QALY (Chinese utilities), obtained under a median-anchored, maturity-constrained lognormal control-arm OS curve calibrated to the published PROpel BRCAm control-arm median OS of 23.0 months (§2.5). The BRCAm result was stable when we propagated the full published OS-HR confidence interval (0.14-0.56) (ICER CNY 53,531-62,444/QALY, §4.3) and when we truncated the modelled intervention OS median from about 105 months down to 29-37 months against external MAGNITUDE anchors (ICER CNY 53,328-53,808/ QALY, §4.7), showing that long-tail extrapolation does not drive the BRCAm conclusion. Both populations remained cost-effective under a two-transition Markov structural specification (ITT co-primary ICER CNY 80,479/QALY, 2.00× the PSM; BRCAm CNY 81,878/QALY, 1.50×; BRCAm × Xu CNY 120,558/QALY, 1.77×).

Under probabilistic sensitivity analysis (5,000 iterations) the probability of cost-effectiveness at the 3× GDP threshold was ≥99.9% in every scenario. The maximum ICER across all structural, utility and population scenarios (BRCAm × Xu Markov, CNY 120,558/QALY) remains 42% of the WTP threshold, so the cost-effectiveness conclusion is invariant to structural, utility and population choice.

### 5.2 Comparison with previous studies

The earlier Chinese CUA of olaparib monotherapy based on the PROfound trial reported an ICER of CNY 392,728/QALY and concluded that olaparib was not cost-effective in China.^22^ Three factors drive our substantially more favourable result. First, and quantitatively dominant, the eleventh round of national VBP reduced olaparib prices by more than 98%, from about CNY 12,240 to CNY 188.7 per cycle,^17^ compressing the incremental drug-cost component from more than CNY 12,000 per cycle to about CNY 188.7 per cycle. Second, the BRCA1/2-confirmed subgroup analysed here shows much larger survival benefit than the PROfound population (PROpel BRCAm rPFS HR 0.23, OS HR 0.29 vs PROfound rPFS HR 0.34, OS HR 0.69)^9,7,38–39^; the FDA’s PARP-inhibitor-by-gene pooled analysis confirms that BRCA1/2-altered patients derive the largest benefit across trials.^8^ Third, our base case uses Castro 2024 pooled utilities^25^ rather than the second-line Xu 2022 values^22^; using the Xu values in a scenario analysis narrows incremental QALY gain from 3.55 to 2.86 and raises the ICER to CNY 68,000/QALY, still highly cost-effective. The BRCAm survival benefit and the utility choice each move the ICER within the same order of magnitude, whereas the VBP price change alone accounts for a factor of order 10 on the incremental drug-cost component; the three factors are therefore not equal contributors. A useful counterfactual would re-run the same PROpel BRCAm model at pre-VBP olaparib pricing (CNY 12,240 per cycle rather than CNY 188.7). The incremental drug-cost differential would then increase by more than an order of magnitude on that component, and the base-case ICER would move materially closer to or above the WTP threshold. The qualitative “cost-effective under some Chinese WTP anchors” conclusion could nonetheless shift depending on the exact scenario. This counterfactual reinforces that the VBP-induced price reset is the operative driver rather than a marginal one.

The results align with a second, contemporaneous Chinese CUA. Pang et al. 2023 constructed a three-state PSM of second-line olaparib monotherapy vs enzalutamide in BRCA1/2-altered mCRPC using PROfound BRCA subgroup efficacy and Chinese unit costs.^53^ Their base case gave incremental cost CNY 48,587, incremental QALY 0.5647, ICER CNY 86,043/QALY, below the 2022 3× GDP threshold of CNY 257,094/QALY. The two analyses are not directly comparable in point-estimate ICER. Pang used a second-line PROfound population (rPFS HR 0.34, OS HR 0.63) with monthly cycles and a 10-year horizon, whereas we use the first-line PROpel BRCAm subgroup (rPFS HR 0.23, OS HR 0.29) with the same cycle length and model horizon (§2.2). Nonetheless, the qualitative agreement across two independent Chinese analyses of PARP inhibition in BRCA1/2-mutated mCRPC supports the value proposition.

The primary contribution of this work is methodological. We applied four pre-specified methodological refinements to a Chinese first-line BRCAm mCRPC CUA under post-VBP pricing. First, we used Guyot pseudo-IPD reconstruction from a published Number-at-Risk table as the input to parametric distributional sensitivity, which removes the circularity of fitting alternative distributions to pseudo-IPD simulated under the base-case assumption. Second, we performed per-scenario PSM cycle-crossing quantification to make the PD-clipping artefact legible when the HR_PFS < HR_OS asymmetry drives S_int_PFS(t) > S_int_OS(t) at long horizons. Third, we applied an HR bound sensitivity (HR_OS = HR_PFS) to quantify the ICER impact of the OS PH assumption. Fourth, we conducted a multi-metamodel EVPPI comparison (§2.6) to test variance-attribution rankings against metamodel choice. The set is portable to any prior CUA with a published Number-at-Risk table, a PSM or Markov structure with cycle-crossing potential, HR-based scaling between arms and a PSA-based decision-uncertainty analysis.

The findings are consistent with recent Chinese CUAs of other advanced prostate cancer treatments. Zhu et al. 2025 reported that talazoparib plus enzalutamide at pre-VBP prices was not cost-effective, with talazoparib drug cost as the dominant sensitivity driver,^40^ reinforcing that PARP-inhibitor combinations require substantial price reductions to reach Chinese WTP thresholds — precisely what the eleventh-round VBP has delivered for olaparib. Chinese CUAs of second-generation androgen-receptor antagonists in mHSPC have concluded cost-effectiveness at similar WTP thresholds,^41–42^ and a US-perspective HRR-guided decision analysis of talazoparib plus enzalutamide by Rui et al. 2025 reported ICER-to-WTP ratios comparable to those observed in the Chinese cost environment.^54^

### 5.3 Trial-level context and post-progression landscape

Three phase-3 competitors form the immediate clinical context for PROpel. MAGNITUDE (niraparib plus abiraterone in HRR-selected patients) showed a comparable BRCAm rPFS benefit (rPFS HR 0.53, 95% CI 0.36-0.79).^11–12^ The final OS analysis at median follow-up 37.3 months reported observed median OS in the BRCA1/2 subgroup of 30.36 months with niraparib plus abiraterone vs 28.55 months with placebo plus abiraterone (unadjusted HR 0.788, 95% CI 0.554-1.120; IPCW-adjusted HR 0.645, 95% CI 0.415-1.002 after correction for imbalanced subsequent therapy).^55^ TALAPRO-2 (talazoparib plus enzalutamide, unselected) reported final OS benefit in the ITT cohort (observed median 45.8 vs 37.0 months; HR 0.80, 95% CI 0.66-0.96) at median follow-up 52.5 months,^15^ with HRR-deficient subgroup rPFS HR 0.45 (95% CI 0.33-0.61)^14^ and ITT rPFS HR 0.63 (95% CI 0.51-0.78).^13^ PROfound established olaparib monotherapy activity in HRR-altered mCRPC after progression on an androgen-receptor pathway inhibitor, with a mature BRCA1/2 subgroup OS HR of 0.63 (95% CI 0.42-0.95).^38–39^ The FDA pooled analysis by individual gene^8^ confirms that BRCA1/2-altered patients derive the largest treatment effect across trials, supporting the BRCAm-focused base case.

The PROpel BRCAm subgroup carries the strongest reported OS signal among these first-line PARPi trials, and this is the single most important interpretive caveat for the present analysis. The final descriptive OS hazard ratio in the PROpel BRCA1/2 subgroup was 0.29 (95% CI 0.14-0.56),^9,7^ well below the MAGNITUDE BRCA1/2 estimates (unadjusted 0.788; IPCW-adjusted 0.645)^55^ and the TALAPRO-2 ITT estimate (0.80).^15^ Applied through proportional-hazards scaling to the median-anchored control curve (median OS 23.0 months) over a 15-year horizon, this HR yields a modelled intervention-arm OS median of about 105 months — roughly 3.5× the MAGNITUDE BRCA1/2 niraparib observed median (30.36 months)^55^ and about 2.3× the TALAPRO-2 ITT talazoparib observed median (45.8 months).^15^ In other words, the modelled BRCAm intervention survival lies beyond the optimistic extreme of the observed first-line PARPi evidence base rather than within it, for two compounding reasons that we state plainly. First, the true PROpel BRCAm OS benefit is genuinely large but a small subgroup (85 patients; 47 olaparib plus abiraterone with 13 deaths, 38 placebo plus abiraterone with 25 deaths) informs it, so the point HR of 0.29 carries wide statistical uncertainty (95% CI 0.14-0.56) and the intervention-arm median was not reached at data cut-off.^9,7^ Second, because the control-arm data cut-off is about 36.6 months, we extrapolate the intervention curve over roughly nine additional years as a parametric projection well beyond observed follow-up. We address both features quantitatively below (external-HR scenario) and in §5.5(2) (probabilistic propagation of the full HR confidence interval). The Chinese NMPA approval on 31 July 2025 was granted for the BRCAm indication only,^16^ mirroring the FDA labelling and reflecting the concentration of the strongest PROpel effect in the BRCA1/2-altered subgroup.^8^

To bound the effect of this strong-OS-benefit extrapolation against external evidence, we ran a conservative scenario analysis substituting the MAGNITUDE final BRCA1/2 OS hazard ratio for the PROpel BRCAm OS HR of 0.29 while holding the PROpel BRCAm rPFS HR (0.23) and all other inputs at their base-case values. This scenario deliberately replaces the strongest observed OS signal (PROpel HR 0.29) with a substantially weaker external one to test whether the cost-effectiveness conclusion depends on the large PROpel BRCAm benefit. Under the MAGNITUDE unadjusted OS HR of 0.788 (95% CI 0.554-1.120)^55^ the modelled intervention-arm OS median falls from about 105 months to about 29 months. The BRCAm ICER was then CNY 53,328/QALY under Castro utilities and 65,741/ QALY under Xu utilities. Under the IPCW-adjusted MAGNITUDE OS HR of 0.645 (95% CI 0.415-1.002)^55^ the modelled intervention median was about 37 months, and the corresponding ICERs were CNY 53,432 and 65,908/QALY. The direction of the effect is initially counter-intuitive and warrants explanation: imposing the weaker MAGNITUDE OS benefit lowers the ICER slightly (by about 2-3% under either utility source) rather than raising it. The mechanism is that a weaker OS benefit shortens post-progression (PD-state) survival in the intervention arm. This reduces the accrual of the relatively costly PD-state management over the horizon. The incremental lifetime cost falls from about CNY 194,000 to about CNY 185,000, while the incremental QALY falls only modestly (from 3.55 to about 3.46 under Castro), so the ratio declines. The key inference is one of robustness rather than convergence: the PROpel base case is driven by an OS benefit considerably stronger than the external MAGNITUDE benchmark, yet substituting the weaker external signal leaves the combination highly cost-effective (about 19-23% of the WTP threshold under both utility sources). The cost-effectiveness conclusion therefore does not depend on accepting the full magnitude of the PROpel BRCAm OS benefit; it holds even under the more conservative externally observed OS hazard ratios.

The study evaluates the first-line indication and does not explicitly model subsequent-line therapy, although the PD-state cost captures post-progression drug and supportive costs. The therapeutic landscape shifted recently with the NMPA approval on 5 November 2025 of lutetium-177 vipivotide tetraxetan (Pluvicto) for PSMA-positive mCRPC after an androgen-receptor pathway inhibitor and optionally taxane chemotherapy.^20–21^ Because Pluvicto is a high-cost option and post-first-line eligibility depends on the PFS duration under first-line therapy, the present PSM may underestimate the sequential economic value of first-line olaparib plus abiraterone (which extends PFS by more than a factor of three in the BRCAm setting under our extrapolation). Future work should model the sequence of first-line PARP inhibitor plus androgen-receptor pathway inhibitor followed by later-line PSMA-targeted radioligand therapy.

### 5.4 Policy implications

The results support inclusion of olaparib plus abiraterone in China’s National Reimbursement Drug List for BRCAm mCRPC. Cumulative 5-year budget impact is modest under all uptake trajectories (deterministic CNY 24.03 million to CNY 75.53 million; probabilistic base-uptake mean CNY 42.42 million), and the BRCA-testing prerequisite enables targeted allocation of scarce resources.

The cost-effectiveness conclusion holds across the Chinese WTP anchors used in reimbursement and provincial VBP negotiations. The base-case ICER of CNY 54,708/QALY represents 57.1% of 1× GDP (CNY 95,749), 38.1% of 1.5× GDP (CNY 143,624), 28.6% of 2× GDP (CNY 191,498), and 19.0% of 3× GDP (CNY 287,247). Probability of cost-effectiveness for BRCAm × Castro (Supplementary Table S2 Panel A) was 99.8% at 1× GDP under both PH scaling (Path A) and AFT non-PH sensitivity (Path B); both paths reached 100% at 1.5× GDP and above. The base-case ITT ICER (CNY 40,198/QALY) suggests broader use may also be economically justified,^24^ although the current NMPA indication is restricted to BRCAm.^16^ The BRCAm × Xu ICER (CNY 68,000/QALY, 23.7% of 3× GDP) is an independent upper bound derived from a Chinese-anchored utility source; because Xu 2022 sampled a second-line population, this scenario provides a conservative ceiling for first-line utility.

Incremental QALYs are invariant to unit drug price by construction (state occupancy and utilities do not depend on drug cost per §4.4), so the ICER shifts linearly with olaparib price via the drug-cost-differential term. Solving deterministically for the olaparib per-cycle price at which the BRCAm base-case ICER equals four common WTP anchors gives ceilings of CNY 8,797 (Castro) and CNY 6,718 (Xu) at 3× GDP (46.6× and 35.6× the current post-VBP price of CNY 188.7). The corresponding ceilings are CNY 5,252/3,867 at 2× GDP, CNY 3,480/2,441 at 1.5× GDP, and CNY 1,708/1,015 at 1× GDP (9.1× and 5.4×). Under an ITT-scenario ceiling recomputation with HR_PFS 0.66, HR_OS 0.81, the ITT × Castro ceiling is CNY 3,159 (16.7×) at 3× GDP and CNY 857 (4.5×) at 1× GDP; ITT × Xu is CNY 2,881 (15.3×) and CNY 764 (4.0×). The table below gives the full 16-cell headroom grid.

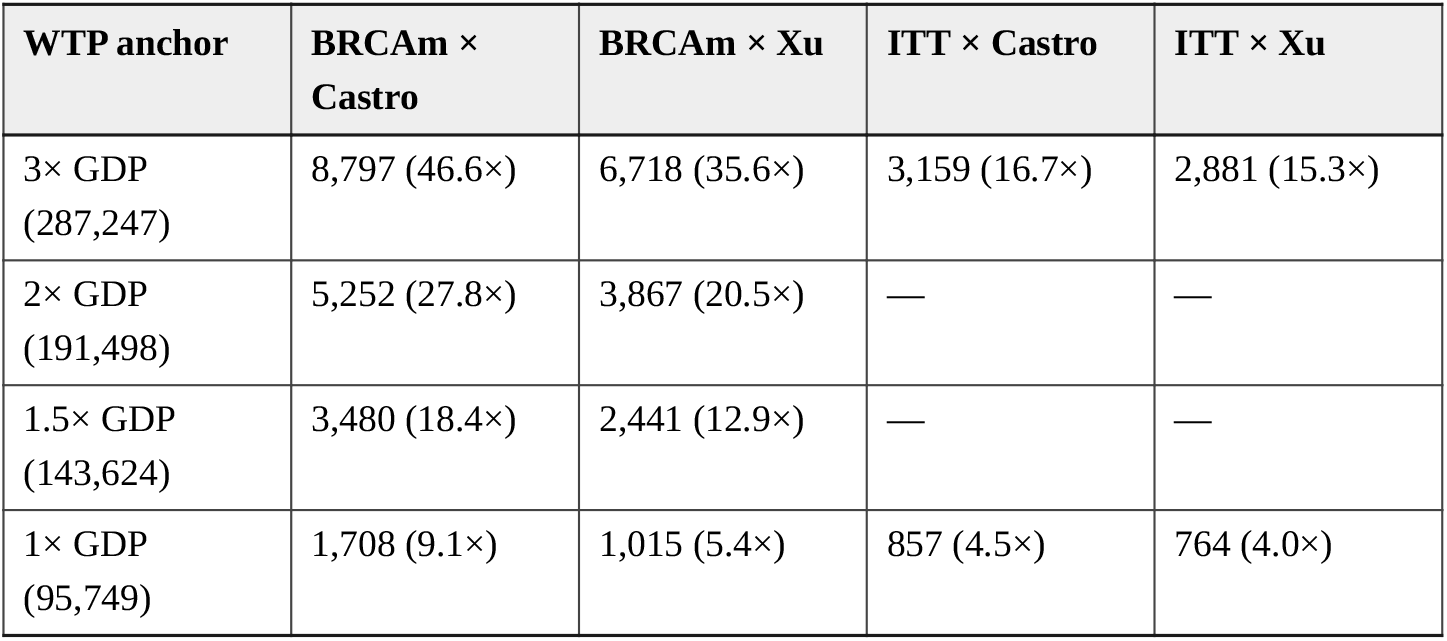

| WTP anchor | BRCaM × Castro | BRCaM × Xu | ITT × Castro | ITT × Xu |
| --- | --- | --- | --- | --- |
| 3× GDP<br>(287,247) | 8,797 (46.6×) | 6,718 (35.6×) | 3,159 (16.7×) | 2,881 (15.3×) |
| 2× GDP<br>(191,498) | 5,252 (27.8×) | 3,867 (20.5×) | — | — |
| 1.5× GDP<br>(143,624) | 3,480 (18.4×) | 2,441 (12.9×) | — | — |
| 1× GDP<br>(95,749) | 1,708 (9.1×) | 1,015 (5.4×) | 857 (4.5×) | 764 (4.0×) |

Prices in CNY per cycle; multiples are relative to the current post-VBP price of CNY 188.7. We omit the ITT 2× GDP and 1.5× GDP anchors as intermediate and not policy-active. These headroom multiples confirm that the current post-VBP olaparib price sits well below the WTP-anchored ceiling under every common Chinese threshold and define a bound above which future price adjustments would erode the cost-effectiveness case. The ITT ceilings remain above the current post-VBP price at all four anchors, so even under an extended reimbursement scope beyond the current BRCAm restriction the current price would remain cost-effective. This ITT conclusion is conditional on the PROpel-reported OS hazard ratio of 0.81; under the more conservative HR_OS = HR_PFS bound of §4.3, ITT × Castro rises to CNY 71,652 and ITT × Xu to CNY 95,872, which remain below the WTP threshold but shift ITT × Castro to about 25% of WTP (from 14% under the base case).

BRCA testing capacity building for tier-2 and tier-3 hospitals should be a prerequisite condition of NRDL listing for olaparib plus abiraterone, or at minimum a co-implementation policy commitment. The 30% to 70% BRCA testing ramp used in the BIA (§3.6) is a policy scenario reflecting present-day inter-provincial heterogeneity, not an empirical forecast. Its upper bound (70% by year 5) is conditional on the co-implementation package being in place. Real-world testing uptake should be tracked prospectively and the BIA re-run if uptake trajectories diverge materially from the assumed ramp. Two prevalence caveats are relevant to the BIA denominator. The BRCA1/2 prevalence of 10% used in the BIA cascade reflects Western trial-population estimates^7–8^; an unselected Chinese cohort reported germline BRCA1/2 mutations in 5.6% of prostate cancer patients,^5^ and disease-stage-conditional prevalence is expected to be higher because BRCAm patients are enriched at advanced stages.^3^ The eligible BRCAm-identified population in the base-case BIA is therefore an upper-envelope estimate; if disease-stage-conditional Chinese prevalence data become available, the BIA should be recomputed with the empirical share. We did not conduct a formal distributional cost-effectiveness analysis (DCEA)^56–57^ quantifying the differential impact of NRDL listing across Chinese provincial income deciles, urban-rural strata, or ethnic minority populations (CHEERS 2022 Item 19); this is a methodological gap for equity-sensitive reimbursement in a country with wide inter-provincial GDP variation and we propose it as future work. The modelling approach follows the Chinese Guidelines for Pharmacoeconomic Evaluations,^23,43^ ISPOR-SMDM good research practice^26–28,35^ and CHEERS 2022 reporting standards.^34^

### 5.5 Strengths, limitations and generalisability

Strengths. The analysis combines three methodological improvements over prior Chinese olaparib mCRPC CUAs: parametric survival modelling on Guyot pseudo-IPD with distribution selection guided by AIC/BIC^24,29–30^ rather than assumed Weibull shape; explicit structural comparison between PSM and Markov specifications, per ISPOR-SMDM good research practice for structural uncertainty^27^; and a full value-of-information analysis including per-patient and population EVPI^31–32^ and nonparametric EVPPI.^33^ Dual utility sourcing tests sensitivity to the well-known tension between global and Chinese utility values in mCRPC.^22,25^ We took adverse-event rates directly from the PROpel safety population without bracketing framing.^7^

Limitations. Eleven limitations warrant acknowledgement, of which the second is the principal driver of residual uncertainty. Several analyses reported here were post hoc (§2.2); we flag them at first mention and their results should be read as supportive rather than confirmatory. (1) Structural uncertainty between PSM and Markov. Two-transition Markov ICERs are 50-77% higher than PSM in the BRCAm subgroup and 100-140% higher in the ITT population under the same lognormal survival parametrisation, reflecting genuine structural uncertainty in long-tail post-progression survival modelling^58–59,27,60^; both structures give cost-effective results at the WTP threshold (maximum ICER 42% of WTP), but the gap cautions against over-precision. The gap is larger under the true BRCAm parameters than in earlier specifications because the very strong OS benefit (HR 0.29) produces a long intervention-arm post-progression occupancy in the Markov structure (§3.2), over which PD-state management cost accrues. A follow-up analysis (§3.2) replaced the constant Markov PD→Death hazard with Weibull (κ=1.5) and Gompertz (γ=0.02/mo) alternatives in the ITT population (the single-transition BRCAm variant is undefined because no non-negative PD→Death hazard can reproduce the prespecified 79.3-month OS target from the PH-scaled rPFS curve, so calibration reaches the zero-hazard boundary). The resulting ITT Markov ICERs moved by 4-7% relative to the constant-hazard Markov baseline. This confirms that the structural constraint of an exponential PD-state exit, rather than the functional form of the PD→Death hazard, drives the PSM-vs-Markov gap. (2) BRCAm intervention-arm extrapolation beyond trial follow-up (principal driver). Under proportional-hazards scaling of the median-anchored control curve (median OS 23.0 months) by the final descriptive OS HR of 0.29, the modelled BRCAm intervention-arm OS median is about 105 months. This substantially exceeds both the PROpel data cut-off of about 36.6 months^7^ and the observed cross-trial first-line PARPi medians (§5.3). We therefore extrapolate the intervention curve over roughly nine years beyond observed follow-up as a lognormal projection. This large extrapolation reflects a genuinely strong but imprecisely estimated subgroup OS benefit: the PROpel BRCAm OS HR of 0.29 is based on only 85 patients (13 deaths of 47 on olaparib plus abiraterone; 25 deaths of 38 on placebo plus abiraterone) and carries a wide 95% CI (0.14-0.56).^9,7^ Two features of the analysis contain the resulting uncertainty. First, we did not fit the control-arm curve to the sparse BRCAm placebo Kaplan-Meier data but anchored it to the single published control-arm median (23.0 months) and constrained it to reproduce the observed control-arm mortality at cut-off (§2.5), which avoids spurious precision from digitising a 38-patient step function (NICE DSU TSD 14).^29^ Second, the PSA propagates the full BRCAm OS HR confidence interval (0.14-0.56); even so, the PSA 97.5th-percentile ICER for BRCAm × Castro (CNY 81,808/QALY) remains about 28% of the WTP threshold, consistent with the 100% probability of cost-effectiveness at 3× GDP in Table 4. The conservative external-HR scenario in §5.3 further shows that substituting the weaker MAGNITUDE OS hazard ratios leaves the combination cost-effective. Readers should nonetheless interpret the base-case CNY 54,708/QALY point estimate as conditional on a strong OS benefit estimated from a small subgroup and extrapolated well beyond trial follow-up; this is the principal source of residual uncertainty in the analysis. (3) Xu 2022 second-line utility applied to first-line population. Applying second-line-derived Xu 2022 utilities^22^ as a first-line scenario introduces conservative bias toward the null (fewer incremental QALYs), quantified in §3.1 against Castro 2024.^25^ (4) Non-proportional hazards in OS. Log-cumulative-hazard plots showed a modest departure from PH for OS (slope ratio 1.19) versus rPFS (slope ratio 1.02); the HR bound sensitivity in §4.3 confirms BRCAm stability (+2.1% to +2.8%, because the true BRCAm OS and rPFS hazard ratios 0.29 and 0.23 are already close) while showing ITT sensitivity (+78.2% to +119.3%). (5) Small BRCAm subgroup (global n=85; Chinese n=15). The PROpel BRCAm subgroup is small, so HRs carry substantial uncertainty; the wide PSA distributions and the 95% uncertainty intervals in Table 4 reflect this. (6) Healthcare payer perspective. We did not include indirect costs (productivity losses, informal caregiver time); a societal perspective would further favour intervention. (7) Line-of-therapy vs disease-state utility mapping. Castro 2024 reported utilities as line-of-therapy values (first-line pooled u=0.79; second-line-and-later 0.69), which we map to PFS and PD respectively^25^; Xu 2022 uses genuinely disease-state Chinese utilities but from a second-line PROfound population.^22^ Neither source is fully aligned with the base-case disease states, so the BRCAm ICER range of CNY 54,708-68,000 across the two sources is a bracketing but incomplete calibration; Chinese first-line disease-state EQ-5D data would further refine utility values. (8) EVPPI subset shares are directional (Strong-Oakley-Brennan). EVPPI subsets are not variance components; each is individually bounded above by total scenario EVPI, but subsets can share information via the metamodel because parameters are jointly informative for the net-monetary-benefit surface. Subset shares therefore sum above 100% (203-219% for RF) as a structural property of the framework^33^ rather than evidence of overfitting; the OOB share differed from the RF share by less than 1 percentage point in 20 of 24 subset-scenario combinations, and the dominant subset was preserved across RF, OOB and GAM in every scenario. (9) Distributional selection is applicable only to ITT. The ITT placebo-arm Guyot pseudo-IPD supports comparison among parametric OS tails; on these data, lognormal (AIC 2014.1) fits less well by AIC than log-logistic (1997.2) and generalised gamma (2000.6), with Δ AIC 17.0 from the best-fitting candidate. We retained lognormal as the ITT base case for clinical plausibility and continuity, while disclosing the complete four-distribution ITT range (Castro CNY 36,765-46,955/QALY; Xu CNY 38,101-53,209/QALY), and every ITT estimate remains below 1× GDP. We report no BRCAm Guyot AIC values or multi-distribution envelope: the true BRCAm placebo arm (n=38, 25 OS events) is too sparse for defensible digitisation and distribution selection, so BRCAm uses only the median-anchored, maturity-constrained base curve and represents parametric uncertainty through the PSA intervals in Table 4 and Figure 5. (10) Time-on-treatment not modelled separately from PFS occupancy. Drug costs accrue over full PFS occupancy, implicitly assuming patients remain on olaparib and abiraterone until radiographic progression. In practice, treatment discontinuation for adverse events, patient preference or non-progression clinical events is non-trivial: the PROpel final safety analysis reported adverse-event-driven treatment discontinuation of 13.8% in the olaparib arm vs 7.8% in the placebo arm at median follow-up 36.6 months.^52^ The base case therefore modestly overstates real-world drug spend and biases the ICER upward relative to a time-on-treatment-modelled counterfactual. The qualitative conclusion of cost-effectiveness is thus conservative under real-world discontinuation patterns. §4.5 now quantifies this: applying treatment-duration multipliers of 0.85 and 0.70 lowers the ITT × Castro ICER from CNY 40,198 to CNY 28,844 and CNY 17,490/QALY, and the BRCAm × Castro ICER from CNY 54,708 to CNY 51,020 and CNY 47,332/QALY. Relaxing the treatment-until-progression assumption therefore improves cost-effectiveness in every scenario. A time-on-treatment reparametrisation using Chinese real-world discontinuation data would refine the point estimate in the favourable direction. (11) PD-state occupancy clipping in the PSM. The partitioned survival cycle loop uses the standard clipped-subtraction identity p_pd = max(S_OS − S_PFS, 0), which forces post-progression occupancy to zero whenever the PH-scaled intervention S_OS(t) falls below its S_PFS(t). As quantified in §4.2, this clipping is active in about 80% of horizon cycles for the BRCAm intervention arm. This is because HR_PFS 0.23 is smaller than HR_OS 0.29, and the PH-scaled intervention-arm OS median (about 105 months) produces a long OS tail relative to the shorter rPFS tail. This is a recognised structural feature of the partitioned survival approach^29,27^ rather than a data artefact. We make its two directional consequences explicit here. First, it makes the BRCAm incremental QALY almost entirely a function of pre-progression (PFS) time, so post-progression utility assumptions have minimal leverage on the BRCAm ICER. Second, it means the BRCAm intervention curve relies on independent PFS and OS extrapolations whose internal consistency is imposed rather than jointly estimated. This clipping is confined to the supporting BRCAm scenario: the primary ITT base case has zero cycle-crossings (§4.2), so the headline ITT conclusion does not depend on the clipped-subtraction identity at all. The Markov structural comparison in §3.2, which does not clip, and the HR-bound analysis in §4.3, which removes the HR_PFS/HR_OS asymmetry, together bound the impact of this feature on the BRCAm scenario; both leave the BRCAm cost-effectiveness conclusion unchanged.

Two operational BIA assumptions are conservative but tentative: the 30-70% BRCA testing ramp over 2025-2029, and the 60% eligibility fraction for first-line combination therapy in Chinese mCRPC. We flag both in Table 6 for adjustment against emerging real-world data.

Generalisability. The base case reflects the BRCAm subgroup that the NMPA indication targets, using global PROpel evidence transported to Chinese unit costs and Chinese utility scenarios. Generalisability to other Asian healthcare systems will depend on drug pricing, BRCA testing capacity and post-progression treatment mix. The ≥99.9% probability of cost-effectiveness at the WTP threshold, stability across two utility sources and two model structures, and zero EVPI at the WTP threshold together indicate that the conclusion is stable under plausible parameter variation. Extension to non-Asian settings will require re-parametrisation of drug prices, PFS management costs and BRCA testing capacity, but we expect the qualitative conclusion (that a BRCA-guided PARP inhibitor combination is economically attractive when drug prices are competitive) to be preserved.

## 6. Conclusion

At current post-VBP prices, first-line olaparib plus abiraterone is cost-effective for first-line mCRPC in China. The primary intention-to-treat result (anchored to the larger PROpel evidence base) gives an ICER of CNY 40,198/QALY (meta-analytic utilities) to CNY 43,721/QALY (Chinese utilities), below 1× the 2024 per-capita GDP, and is robust across four survival distributions (CNY 35,922-53,209/QALY), a ±0.05 utility swing, time-on-treatment reparametrisation, and a two-transition Markov structure; it carries no partitioned-survival clipping (zero cycle-crossings). The prespecified BRCA1/2-mutated subgroup, reported as a supporting precision-medicine analysis consistent with the NMPA-approved indication, shows an at-least-as-favourable signal (ICER CNY 54,708-68,000/QALY) that remains cost-effective across the full published OS-HR confidence interval (0.14-0.56), under external-HR (MAGNITUDE) and OS-cap scenarios truncating the modelled median to as low as 29 months, and under the Markov structure (maximum ICER 42% of WTP). The principal residual uncertainty is specific to the supporting BRCAm analysis: a strong but imprecisely estimated OS benefit (HR 0.29, 95% CI 0.14-0.56) from a small subgroup (n=85, intervention-arm median OS not reached) extrapolated beyond trial follow-up. The sensitivity analyses above bound rather than eliminate this limitation, and it does not affect the primary ITT conclusion; readers should interpret the BRCAm ICER as a bounded, hypothesis-consistent signal rather than a precise estimate. The probability of cost-effectiveness at the WTP threshold is ≥99.9% in every scenario, and per-patient EVPI is zero. The 5-year payer budget impact is modest (deterministic CNY 24.03 million to CNY 75.53 million under low and high uptake; probabilistic base-uptake mean CNY 42.42 million, 95% interval 25.51-64.67 million), with BRCA1/2 prevalence the dominant budget driver. These findings support inclusion of olaparib plus abiraterone in the NRDL, particularly for the NMPA-approved BRCAm indication, together with parallel investment in tier-2 and tier-3 hospital BRCA testing capacity.

## Abbreviations

AFT: Accelerated failure time
AIC: Akaike Information Criterion
BIA: Budget impact analysis
BIC: Bayesian Information Criterion
BRCAm: BRCA1/2-mutated
CHEERS: Consolidated Health Economic Evaluation Reporting Standards
CI: Confidence interval
CNY: Chinese Yuan (Renminbi)
CrI: Credible interval
CUA: Cost-utility analysis
DSU: Decision Support Unit (NICE)
EQ-5D: EuroQol 5-Dimension health-utility instrument
EVPI: Expected value of perfect information
EVPPI: Expected value of partial perfect information
FDA: US Food and Drug Administration
GAM: Generalised additive model
GDP: Gross domestic product
GG: Generalised gamma
HR: Hazard ratio
HRR: Homologous recombination repair
ICER: Incremental cost-effectiveness ratio
IPCW: Inverse probability of censoring weighting
IPD: Individual patient data
ISPOR: Professional Society for Health Economics and Outcomes Research
ITT: Intention-to-treat
KM: Kaplan-Meier
mCRPC: Metastatic castration-resistant prostate cancer
NICE: National Institute for Health and Care Excellence
NMPA: National Medical Products Administration (China)
NRDL: National Reimbursement Drug List (China)
OOB: Out-of-bag
OS: Overall survival
PARP: Poly(ADP-ribose) polymerase
PD: Progressed disease (post-progression health state)
PFS: Progression-free survival
PH: Proportional hazards
PSA: Probabilistic sensitivity analysis
PSM: Partitioned survival model
PSMA: Prostate-specific membrane antigen
QALY: Quality-adjusted life-year
rPFS: Radiographic progression-free survival
SMDM: Society for Medical Decision Making
TSD: Technical Support Document (NICE DSU)
USD: United States Dollar
VBP: Volume-based procurement
WTP: Willingness-to-pay

## Ethical approval

This study did not involve human or animal subjects and therefore did not require ethical approval.

## Consent

This study did not involve patient data, and no consent was required.

## Author contributions

H.D., L.J., Q.T.: Writing – original draft, writing – review and editing, and software. C.Y.: Software. D.Y.: Data curation. A.C.: Data curation. J.X.: Data curation. H.X.: Formal analysis. N.Z.: Investigation. B.Z.: Methodology. M.F.: Resources, supervision, and visualization. J.S.: Conceptualization and writing – original draft.

## Reporting checklist

CHEERS 2022 checklist: provided as Supplementary Material.^34^

## Data Availability

All model input data are provided in Table 1 and the supplementary material of the manuscript. The reconstructed pseudo-individual patient data, full model outputs, and the Python source code that regenerates all base-case results will be made publicly available upon publication.

---

**Supplementary Table S4.**
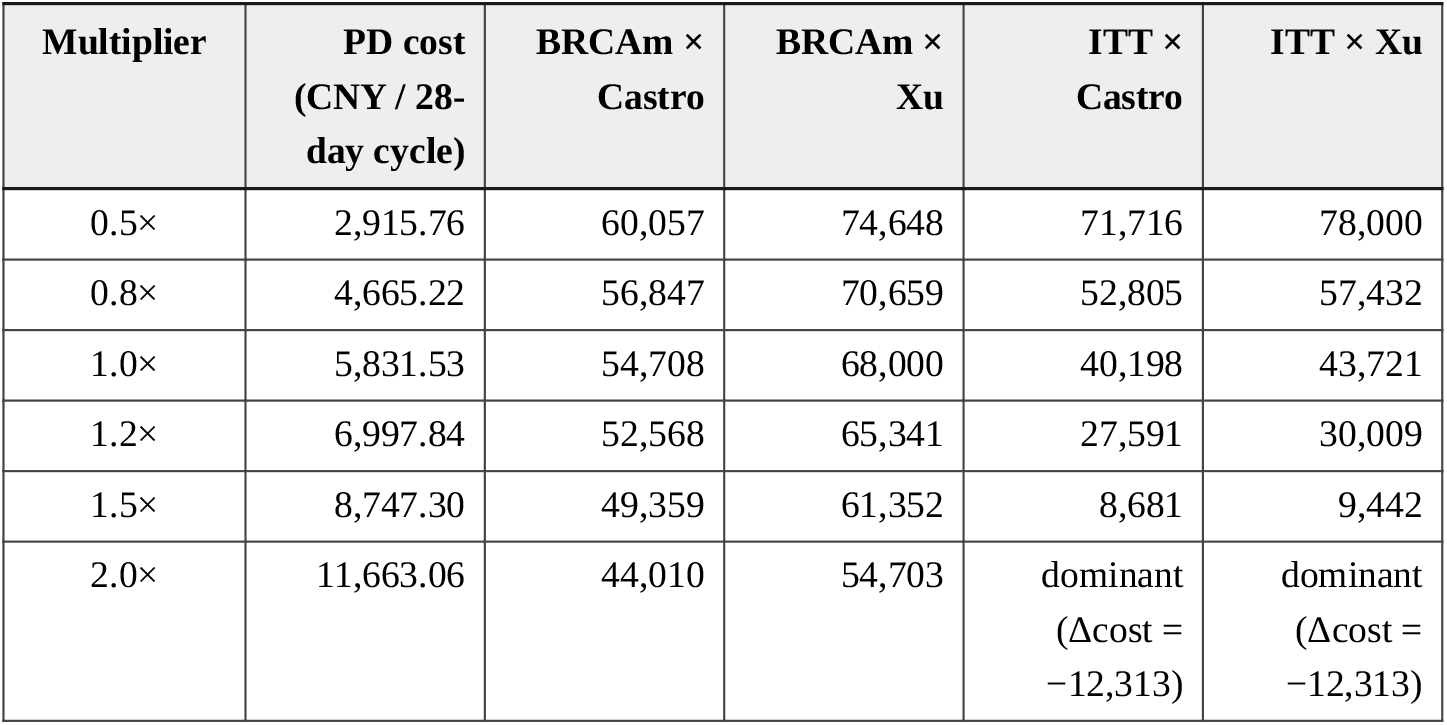
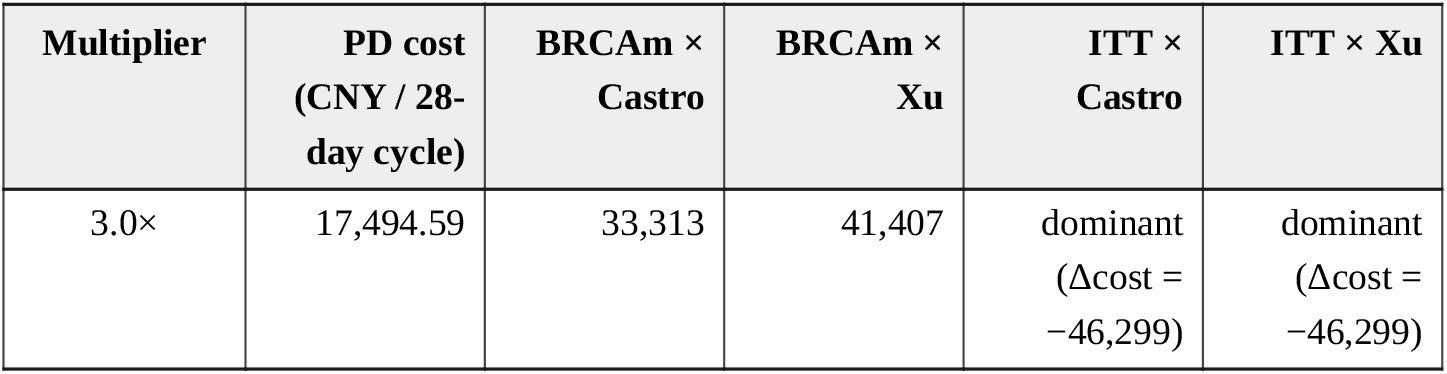
PD-state cost multiplier sensitivity (7 multipliers × 4 published scenarios; ICER in CNY/QALY; deterministic re-scoring at base-case values for all other parameters). BRCAm columns use the true-BRCAm median-anchored control curve and HR_OS 0.29; ITT columns are unaffected by the subgroup and are carried forward unchanged.

