## Supplementary Tables S1-S9 for "Cost-Utility Analysis of First-Line Olaparib plus Abiraterone for Metastatic Castration-Resistant Prostate Cancer in China after Volume-Based Procurement"

### Supplementary Table S1. EVPPI metamodel robustness comparison

| Scenario | Subset | RF in-sample (CNY, %) | RF out-of-bag (CNY, %) | GAM (CNY, %) | RF vs OOB Δ (pp) |
| --- | --- | --- | --- | --- | --- |
| BRCAm × Castro | HRs (rPFS+OS) | 5,477 (34.2%) | 5,458 (34.1%) | 5,414 (33.8%) | 0.12 |
| BRCAm × Castro | Utilities | 4,740 (29.6%) | 4,698 (29.4%) | 4,056 (25.3%) | 0.27 |
| BRCAm × Castro | Drug costs (ola+abi) | 4,341 (27.1%) | 4,299 (26.9%) | 3,751 (23.4%) | 0.26 |
| BRCAm × Castro | PFS mgmt cost | 13,330 (83.3%) | 13,342 (83.4%) | 13,164 (82.3%) | 0.07 |
| BRCAm × Castro | PD cost | 3,632 (22.7%) | 3,635 (22.7%) | 1,354 (8.5%) | 0.02 |
| BRCAm × Castro | AE rates + disutilities | 2,439 (15.2%) | 2,110 (13.2%) | 1,461 (9.1%) | 2.05 |
| BRCAm × Xu | HRs (rPFS+OS) | 6,877 (37.9%) | 6,830 (37.7%) | 6,862 (37.8%) | 0.26 |
| BRCAm × Xu | Utilities | 8,282 (45.7%) | 8,232 (45.4%) | 8,098 (44.6%) | 0.27 |
| BRCAm × Xu | Drug costs (ola+abi) | 4,121 (22.7%) | 4,087 (22.5%) | 3,394 (18.7%) | 0.19 |
| BRCAm × Xu | PFS mgmt cost | 13,012 (71.7%) | 13,041 (71.9%) | 12,702 (70.0%) | 0.16 |
| BRCAm × Xu | PD cost | 3,622 (20.0%) | 3,645 (20.1%) | 1,020 (5.6%) | 0.13 |
| BRCAm × Xu | AE rates + disutilities | 2,196 (12.1%) | 1,767 (9.7%) | 978 (5.4%) | 2.36 |
| ITT × Castro | HRs (rPFS+OS) | 6,922 (82.9%) | 6,899 (82.6%) | 7,067 (84.6%) | 0.27 |
| ITT × Castro | Utilities | 1,467 (17.6%) | 1,427 (17.1%) | 696 (8.3%) | 0.48 |
| ITT × Castro | Drug costs (ola+abi) | 1,660 (19.9%) | 1,628 (19.5%) | 1,118 (13.4%) | 0.38 |
| ITT × Castro | PFS mgmt cost | 3,367 (40.3%) | 3,376 (40.4%) | 2,895 (34.7%) | 0.11 |
| ITT × Castro | PD cost | 3,021 (36.2%) | 3,026 (36.2%) | 2,658 (31.8%) | 0.06 |
| ITT × Castro | AE rates + disutilities | 1,059 (12.7%) | 867 (10.4%) | 563 (6.7%) | 2.30 |
| ITT × Xu | HRs (rPFS+OS) | 8,419 (86.4%) | 8,378 (86.0%) | 8,591 (88.1%) | 0.41 |
| ITT × Xu | Utilities | 1,841 (18.9%) | 1,794 (18.4%) | 1,085 (11.1%) | 0.49 |
| ITT × Xu | Drug costs (ola+abi) | 1,770 (18.2%) | 1,727 (17.7%) | 1,051 (10.8%) | 0.44 |
| ITT × Xu | PFS mgmt cost | 3,414 (35.0%) | 3,426 (35.2%) | 2,807 (28.8%) | 0.13 |
| ITT × Xu | PD cost | 3,109 (31.9%) | 3,123 (32.0%) | 2,600 (26.7%) | 0.14 |
| ITT × Xu | AE rates + disutilities | 1,189 (12.2%) | 969 (9.9%) | 592 (6.1%) | 2.26 |

Notes: RF = RandomForestRegressor (n_estimators=200, max_features=‘sqrt’, min_samples_leaf=20, seed=20260707); OOB = out-of-bag prediction from the same forest; GAM = LinearGAM (pygam v0.12) with 10 spline basis functions per feature. The BRCAm rows are recomputed on the updated PSA (true BRCAm HR_OS 0.29 / HR_PFS 0.23, seed 20260707, n=5000); the ITT rows use the ITT PSA inputs (HR_OS 0.81 / HR_PFS 0.66) and are therefore unaffected by the BRCAm subgroup parameterisation. Percentages express EVPPI as a share of the scenario peak-EVPI WTP total EVPI (BRCAm × Castro: CNY 16,002 at WTP 57,000/QALY; BRCAm × Xu: CNY 18,139 at WTP 72,000/QALY; ITT × Castro: CNY 8,351 at WTP 41,000/QALY; ITT × Xu: CNY 9,748 at WTP 44,000/QALY). RF vs OOB Δ is the absolute percentage-point difference between RF and OOB EVPPI share (≤0.3 pp for all subsets except the adverse-event subset, where OOB is about 2.1–2.4 pp lower). The dominant subset (row with highest RF %) is preserved across RF, OOB, and GAM in all four scenarios: PFS management cost dominates both BRCAm scenarios; hazard ratios dominate both ITT scenarios.

### Supplementary Table S2. Survival-structure robustness of the base case: proportional-hazards versus accelerated-failure-time PSA (Panel A) and parametric-distribution matrix (Panel B; ITT multi-distribution + BRCAm median-anchored base case)

**Panel A. PSA by hazards structure (BRCAm × Castro, 5,000 iterations, seed 42; true BRCAm HR_OS 0.29)**

| PSA path | Mean ICER (CNY/QALY) | Median ICER (CNY/QALY) | 95% CrI (CNY/QALY) | Mean incremental cost (CNY) | Mean incremental QALY | CE@1×GDP | CE@1.5×GDP | CE@2×GDP | CE@3×GDP |
| --- | --- | --- | --- | --- | --- | --- | --- | --- | --- |
| PH-scaling (Path A) | 56,461 | 56,241 | 40,498 – 73,185 | 198,899 | 3.534 | 99.82% | 100% | 100% | 100% |
| AFT non-PH (Path B) | 56,172 | 56,355 | 41,541 – 69,151 | 195,615 | 3.504 | 99.94% | 100% | 100% | 100% |

Notes (Panel A): Panel A reports two additional 5,000-iteration PSA sensitivity paths (seed 42) on top of the primary PSA (5,000 iterations, seed 20260707) reported in Table 4, computed at the true BRCAm OS HR of 0.29 (the deterministic PH base reproduces the Table 2 BRCAm × Castro ICER of CNY 54,708/QALY exactly). Both paths jointly propagate ITT lognormal maximum-likelihood estimate (MLE) uncertainty into the BRCAm control-arm (μ, σ) via asymptotic bootstrap standard errors, BRCAm hazard ratios via lognormal draws based on the reported 95% CIs (rPFS 0.12–0.43; OS 0.14–0.56), and adverse-event rates via Beta(k, n − k) posteriors; Path A retains the base-case S_int(t) = S_ctrl(t)^HR proportional-hazards structure, while Path B replaces PH-scaling with an accelerated-failure-time structure μ_int = μ_ctrl + log(shift), where shift is drawn from a lognormal with mean 1/HR_OS = 1/0.29 = 3.448 (giving an AFT median of 79.3 months, versus approximately 105.2 months for the PH curve) and 15% coefficient of variation. The proportional-hazards diagnostic flag (PH_break_active = True; observed slope ratio 1.185) was pre-specified to activate Path B when it exceeded 1.15. WTP thresholds: 1×GDP = CNY 95,749; 1.5×GDP = CNY 143,624; 2×GDP = CNY 191,498; 3×GDP = CNY 287,247. The two paths produce mean ICERs within 0.5% of each other (56,461 vs 56,172) despite their different implied medians; this empirical agreement shows that the cost-effectiveness conclusion is robust to the PH-versus-AFT structural choice, without implying that the two survival curves are equivalent.

**Panel B. Parametric-distribution robustness matrix (deterministic base case). ITT: 8 scenarios (2 utility × 4 OS distributions on valid Guyot pseudo-IPD, n=397 placebo, 205 events). BRCAm: median-anchored base case only (the Guyot-fit alternatives are not applicable — see note).**

| Population × utility | OS distribution | Incremental cost (CNY) | Incremental QALY | ICER (CNY/QALY) | Cross I/C |
| --- | --- | --- | --- | --- | --- |
| BRCAm × Castro | Lognormal (median-anchored, base case) | 194,214 | 3.5500 | 54,708 | 156 / 130 |
| BRCAm × Xu | Lognormal (median-anchored, base case) | 194,214 | 2.8561 | 68,000 | 156 / 130 |
| ITT × Castro | Lognormal (median-anchored) | 18,254 | 0.5081 | 35,922 | 0 / 0 |
| ITT × Castro | Lognormal, Guyot (base case) | 21,674 | 0.5392 | 40,198 | 0 / 0 |
| ITT × Castro | Generalised gamma, Guyot | 28,020 | 0.5967 | 46,955 | 125 / 92 |
| ITT × Castro | Log-logistic, Guyot | 18,897 | 0.5140 | 36,765 | 1 / 1 |
| ITT × Xu | Lognormal (median-anchored) | 18,254 | 0.4791 | 38,101 | 0 / 0 |
| ITT × Xu | Lognormal, Guyot (base case) | 21,674 | 0.4957 | 43,721 | 0 / 0 |
| ITT × Xu | Generalised gamma, Guyot | 28,020 | 0.5266 | 53,209 | 125 / 92 |
| ITT × Xu | Log-logistic, Guyot | 18,897 | 0.4822 | 39,187 | 1 / 1 |

Notes: For the ITT population, Panel B computes the deterministic base-case ICER under four alternative overall-survival extrapolation distributions for each of the two utility sets. The three fitted distributions (lognormal base case, generalised gamma, log-logistic) are fitted to Guyot-reconstructed ITT control-arm pseudo-individual patient data (n=397 placebo, 205 events; §2.5) and are ranked by Akaike information criterion in §5.5(9); the “median-anchored” lognormal is an additional reference parameterisation. The ITT Guyot lognormal row reproduces the Table 2 ITT base-case ICERs exactly (ITT × Castro 40,198; ITT × Xu 43,721). The ITT distributional envelope is wide (36,765–46,955 for Castro and 39,187–53,209 for Xu; generalised gamma +16.8% and +21.7%, log-logistic −8.5% and −10.4% relative to the respective lognormal base cases), and all eight ITT scenarios remain below the 1×GDP willingness-to-pay threshold of CNY 95,749/QALY.

For the BRCAm subgroup, the multi-distribution matrix is **not applicable**: the true BRCAm placebo arm (n=38, 25 OS events) is too sparse for Guyot digitisation or discrimination among competing parametric tails without spurious precision (NICE DSU TSD 14) ^29^, so only the median-anchored base case (control OS median 23.0 months; §2.5) is reported. BRCAm parametric uncertainty is instead quantified probabilistically through the PSA (Table 4): the 95% credible interval of the BRCAm × Castro ICER is CNY 37,420–81,808/QALY and of the BRCAm × Xu ICER is CNY 44,747–111,424/QALY (both 100% probability of cost-effectiveness at the WTP threshold), and the PH-versus-AFT structural robustness of the BRCAm base case is shown in Panel A. The BRCAm median-anchored row reproduces the Table 2 BRCAm base-case ICERs exactly (BRCAm × Castro 54,708; BRCAm × Xu 68,000).

“Cross I/C” reports the number of 195-cycle model cycles in which the intervention (I) and control (C) arms had progressed-disease-state occupancy clipped to zero because the proportional-hazards-scaled progression-free survival curve exceeded the overall-survival curve; larger counts indicate more of the horizon spent in the pre-progression state and are a systematic consequence of HR_PFS being smaller than HR_OS (BRCAm 0.23 vs 0.29; §4.2). The ITT distributional matrix is visualised in Figure 6.

### Supplementary Table S3. Budget impact for radioligand therapy Pluvicto (177Lu-vipivotide tetraxetan) in the post-olaparib BRCA1/2-mutated mCRPC pathway — 18-scenario grid

**Base parameters:** annual new mCRPC 25,000 patients (Globocan 2022 China 134,000 prostate-cancer incidence × 18% mCRPC rate); BRCA1/2 prevalence 10%; first-line eligibility 80%; PROpel-based olaparib–abiraterone 5-year model-derived progression rate 32%; PSMA-positive rate 85% (VISION-2 screening); 6 doses per full Pluvicto course. Annual eligible-for-Pluvicto flow = 25,000 × 0.10 × 0.80 × 0.32 × 0.85 × BRCA_test_rate × uptake_rate = 543 × BRCA_test_rate × uptake_rate patients/year.

| BRCA testing scenario | Uptake scenario | Price scenario (CNY / dose) | Eligible patients / year | Annual budget impact | 5-year cumulative budget impact |
| --- | --- | --- | --- | --- | --- |
| Current 2025 (30% test) | Low (5% uptake) | 80,000 | 8 | CNY 3.8 M | CNY 19.2 M |
| Current 2025 (30% test) | Low (5% uptake) | 100,000 | 8 | CNY 4.8 M | CNY 24.0 M |
| Current 2025 (30% test) | Low (5% uptake) | 150,000 | 8 | CNY 7.2 M | CNY 36.0 M |
| Current 2025 (30% test) | Mid (10% uptake) | 80,000 | 16 | CNY 7.7 M | CNY 38.4 M |
| Current 2025 (30% test) | Mid (10% uptake) | 100,000 | 16 | CNY 9.6 M | CNY 48.0 M |
| Current 2025 (30% test) | Mid (10% uptake) | 150,000 | 16 | CNY 14.4 M | CNY 72.0 M |
| Current 2025 (30% test) | High (20% uptake) | 80,000 | 32 | CNY 15.4 M | CNY 76.8 M |
| Current 2025 (30% test) | High (20% uptake) | 100,000 | 32 | CNY 19.2 M | CNY 96.0 M |
| Current 2025 (30% test) | High (20% uptake) | 150,000 | 32 | CNY 28.8 M | CNY 144.0 M |
| Target 2029 (70% test) | Low (5% uptake) | 80,000 | 19 | CNY 9.1 M | CNY 45.6 M |
| Target 2029 (70% test) | Low (5% uptake) | 100,000 | 19 | CNY 11.4 M | CNY 57.0 M |
| Target 2029 (70% test) | Low (5% uptake) | 150,000 | 19 | CNY 17.1 M | CNY 85.5 M |
| Target 2029 (70% test) | Mid (10% uptake) | 80,000 | 38 | CNY 18.2 M | CNY 91.2 M |
| Target 2029 (70% test) | Mid (10% uptake) | 100,000 | 38 | CNY 22.8 M | CNY 114.0 M |
| Target 2029 (70% test) | Mid (10% uptake) | 150,000 | 38 | CNY 34.2 M | CNY 171.0 M |
| Target 2029 (70% test) | High (20% uptake) | 80,000 | 76 | CNY 36.5 M | CNY 182.4 M |
| Target 2029 (70% test) | High (20% uptake) | 100,000 | 76 | CNY 45.6 M | CNY 228.0 M |
| Target 2029 (70% test) | High (20% uptake) | 150,000 | 76 | CNY 68.4 M | CNY 342.0 M |

Notes: Scenarios span 2 BRCA1/2 testing coverage rates (current-2025 30%, target-2029 70%), 3 Pluvicto uptake rates among PSMA-positive post-olaparib patients (low 5%, mid 10%, high 20%), and 3 Pluvicto unit prices (low CNY 80,000, mid CNY 100,000, high CNY 150,000 per dose; each course = 6 doses per VISION protocol). Annual budget impact = eligible patients per year × 6 doses × unit price. The full 18-scenario range spans CNY 3.8 million to CNY 68.4 million per year (annual steady state) and CNY 19.2 million to CNY 342.0 million cumulative over five years. Because the eligible-for-Pluvicto cohort is small in absolute terms (8 to 76 patients per year across the 18-scenario grid), the primary Pluvicto BIA driver is per-dose price (an 87.5% range CNY 80,000 to 150,000 per dose translates directly into the same relative price sensitivity), followed by uptake and then by BRCA testing coverage. The low-corner scenario (CNY 3.8 M annual) is compatible with routine payer accommodation; the high-corner scenario (CNY 68.4 M annual) is approximately 47% higher than the first-line olaparib plus abiraterone base-uptake five-year cumulative BIA of CNY 46.57 million reported in §3.6, and would warrant explicit price negotiation and/or eligibility narrowing prior to VBP inclusion of Pluvicto.

### Supplementary Table S4. PD-state cost multiplier sensitivity

The full PD-state cost multiplier sensitivity table (7 multipliers × 4 published scenarios) is presented with its analytic context in §4.4 to keep the stress-test methodology and results together. There are nine supplementary tables in total (S1, EVPPI metamodel robustness; S2, PSA with ITT-bootstrap propagation and flexsurv distributional robustness; S3, downstream Pluvicto sequencing budget impact; S4, PD-state cost multiplier sensitivity; S5, time-on-treatment sensitivity; S6, one-way utility sensitivity; S7, OS extrapolation cap; S8, budget-impact one-way sensitivity; S9, BRCAm control-curve scale-parameter σ one-way sensitivity) and six main-text tables (Tables 1-6); each is called out at its point of first mention in the main text.

| Multiplier | PD cost (CNY / 28-day cycle) | BRCAm × Castro | BRCAm × Xu | ITT × Castro | ITT × Xu |
| --- | --- | --- | --- | --- | --- |
| 0.5× | 2,915.76 | 60,057 | 74,648 | 71,716 | 78,000 |
| 0.8× | 4,665.22 | 56,847 | 70,659 | 52,805 | 57,432 |
| 1.0× | 5,831.53 | 54,708 | 68,000 | 40,198 | 43,721 |
| 1.2× | 6,997.84 | 52,568 | 65,341 | 27,591 | 30,009 |
| 1.5× | 8,747.30 | 49,359 | 61,352 | 8,681 | 9,442 |
| 2.0× | 11,663.06 | 44,010 | 54,703 | dominant (Δcost = −12,313) | dominant (Δcost = −12,313) |
| 3.0× | 17,494.59 | 33,313 | 41,407 | dominant (Δcost = −46,299) | dominant (Δcost = −46,299) |

### Supplementary Table S5. Time-on-treatment sensitivity (intervention drug-cost multiplier τ; §4.5)

| Scenario | τ = 1.0 (base, treat-to-progression) | τ = 0.85 | τ = 0.70 |
| --- | --- | --- | --- |
| ITT × Castro | 40,198 | 28,844 | 17,490 |
| ITT × Xu | 43,721 | 31,372 | 19,022 |
| BRCAm × Castro | 54,708 | 51,020 | 47,332 |
| BRCAm × Xu | 68,000 | 63,416 | 58,832 |

Notes: ICER in CNY/QALY. τ scales only the incremental intervention drug-cost component (olaparib + abiraterone + prednisone = CNY 910.08/cycle) relative to full pre-progression (PFS) occupancy; τ = 1.0 reproduces the base case (treatment continued until radiographic progression). Because τ < 1 lowers only intervention cost, the ICER falls monotonically as modelled treatment duration shortens, so the base case (τ = 1.0) is the conservative bound on this axis. Every value is below 1× GDP (CNY 95,749/QALY). The PROpel final safety analysis reported adverse-event-driven treatment discontinuation of 13.8% (olaparib arm) vs 7.8% (placebo arm) at median follow-up 36.6 months ^52^, consistent with τ modestly below 1 in practice. Visualised in Figure 9 Panel C.

### Supplementary Table S6. One-way utility sensitivity (±0.05 on each health-state utility; §4.6)

| Scenario | Base | PFS utility +0.05 | PFS utility −0.05 | PD utility +0.05 | PD utility −0.05 | One-way range |
| --- | --- | --- | --- | --- | --- | --- |
| ITT × Castro | 40,198 | 36,546 | 44,661 | 41,936 | 38,599 | 36,546–44,661 |
| ITT × Xu | 43,721 | 39,435 | 49,052 | 45,784 | 41,836 | 39,435–49,052 |
| BRCAm × Castro | 54,708 | 51,153 | 58,794 | 55,095 | 54,326 | 51,153–58,794 |
| BRCAm × Xu | 68,000 | 62,593 | 74,429 | 68,599 | 67,411 | 62,593–74,429 |

Notes: ICER in CNY/QALY. Each PFS and PD utility was varied by ±0.05 around its source-specific central value, one at a time, holding all other inputs at base case. Every one-way perturbation left all four scenarios below the WTP threshold (CNY 287,247/QALY) and preserved the ordering ITT < BRCAm; every ITT value remained below 1× GDP (CNY 95,749/QALY). Visualised in Figure 9 Panel D.

### Supplementary Table S7. OS extrapolation cap — external-anchored survival truncation (BRCAm; §4.7)

| OS truncation anchor | Implied HR_OS | Intervention OS median (months) | BRCAm × Castro | BRCAm × Xu |
| --- | --- | --- | --- | --- |
| Base (PH-scaled) | 0.29 | 105.2 | 54,708 | 68,000 |
| Mid cap (60 months) | 0.4315 | 60.0 | 53,808 | 66,522 |
| MAGNITUDE IPCW-adjusted | 0.645 | 36.7 | 53,432 | 65,908 |
| MAGNITUDE unadjusted | 0.788 | 29.4 | 53,328 | 65,741 |

Notes: ICER in CNY/QALY. The modelled intervention-arm OS median (~105 months under proportional-hazards scaling at HR_OS 0.29) was capped to progressively shorter values by substituting weaker OS hazard ratios drawn from external MAGNITUDE BRCA1/2 anchors ^11-12,55^. The BRCAm ICER changes by < 2.5% even when the intervention median is truncated to 29.4 months, because truncating the low-survival-probability tail removes incremental cost and incremental QALYs roughly proportionally; the BRCAm cost-effectiveness conclusion is therefore not driven by long-tail extrapolation. Visualised in Figure 9 Panel B.

### Supplementary Table S8. Budget-impact one-way sensitivity — 5-year payer budget (§4.8)

| Cascade policy parameter | Range (min–mode–max) | 5-year budget low (M CNY) | 5-year budget high (M CNY) | Swing (M CNY) |
| --- | --- | --- | --- | --- |
| BRCA1/2 prevalence | 0.056–0.10–0.12 | 26.08 | 55.89 | 29.81 |
| Metastatic-disease probability | 0.25–0.30–0.40 | 38.81 | 62.10 | 23.29 |
| Combination-eligibility fraction | 0.45–0.60–0.75 | 34.93 | 58.22 | 23.29 |
| Testing-ramp scale | 0.70–1.15 (uniform) | 32.60 | 53.56 | 20.96 |
| Progression-to-mCRPC probability | 0.50–0.60–0.70 | 38.81 | 54.34 | 15.52 |

Notes: 5-year base-uptake budget = CNY 46.57 million (§3.6). Each cascade policy input was varied one at a time across its PERT/uniform support (all other inputs at base). BRCA1/2 prevalence is the dominant budget driver (swing CNY 29.8 million) — a measurable epidemiological quantity and the highest-value target for Chinese real-world calibration. This one-way tornado uses linear multiplicative scaling from the deterministic base case to rank drivers by magnitude and does not replace the joint probabilistic BIA (§3.6, base-uptake mean CNY 42.42 million, 95% interval 25.51–64.67 million). All one-way excursions remain within a range routinely absorbed in provincial reimbursement. Visualised in Figure 10.

### Supplementary Table S9. BRCAm control-curve scale-parameter (σ) one-way sensitivity (§4.9)

| Scenario | σ − 20% (σ = 0.914) | Base (σ = 1.1422) | σ + 20% (σ = 1.371) | One-way range | Max % of WTP |
| --- | --- | --- | --- | --- | --- |
| BRCAm × Castro | 54,943 | 54,708 | 53,968 | 53,968–54,943 | 19.1% |
| BRCAm × Xu | 68,388 | 68,000 | 66,784 | 66,784–68,388 | 23.8% |

Notes: ICER in CNY/QALY. The scale parameter σ of the BRCAm median-anchored, maturity-constrained lognormal control-arm OS curve was varied ±20% about its solved base-case value (σ = 1.1422), holding the published median (μ = ln 23.0 = 3.1355) and all other inputs at base case; all values recomputed deterministically (§4.9). The ICER range spans at most +0.4% (Castro) and +0.6% (Xu) from base, and the maximum σ-excursion ICER reaches only 19.1% (Castro) and 23.8% (Xu) of the WTP threshold (CNY 287,247/QALY). Data-anchored alternative: re-solving σ to the upper Clopper-Pearson 95% bound of the observed control mortality (0.804, giving σ = 0.544) changed the ICERs by <0.1% (BRCAm × Castro CNY 54,687; BRCAm × Xu CNY 67,965); the lower mortality bound (0.486) is structurally unreachable while holding the published 23.0-month median (§4.9).
