## Supplementary material for "Cost-Utility Analysis of First-Line Olaparib plus Abiraterone for Metastatic Castration-Resistant Prostate Cancer in China after Volume-Based Procurement": Tables 1-6

### Table 1. Model input parameters

| Parameter | Base value | Range / 95% CI | Distribution (PSA) | Source |
| --- | --- | --- | --- | --- |
| **Clinical (ITT)** |  |  |  |  |
| Lognormal rPFS control μ (fitted median 19.82 mo; published 16.6 mo, Saad 2023) | 2.9865 | — | — | Reconstructed KM + MLE ^7,26^ |
| Lognormal rPFS control σ | 1.0453 | — | — | Reconstructed KM + MLE ^7,26^ |
| Lognormal OS control μ (median 36.65 mo) | 3.6013 | — | — | Reconstructed KM + MLE ^7,26^ |
| Lognormal OS control σ | 0.9532 | — | — | Reconstructed KM + MLE ^7,26^ |
| ITT HR rPFS | 0.66 | 0.54–0.81 | Lognormal | ^7^ |
| ITT HR OS | 0.81 | 0.67–1.00 | Lognormal | ^7^ |
| **Clinical (BRCAm, BRCA1/2-confirmed, n=85)** |  |  |  |  |
| Median rPFS control (BRCAm) | 8 months | — | — | ^6^ |
| Median OS control (BRCAm) | 23 months | — | — | ^6,7^ |
| BRCAm HR rPFS | 0.23 | 0.12–0.43 | Lognormal | ^6^ |
| BRCAm HR OS | 0.29 | 0.14–0.56 | Lognormal | ^6,7^ |
| **Utilities (Castro 2024, EQ-5D)** |  |  |  |  |
| PFS utility (1L mapped) | 0.79 | 0.75–0.84 | Beta | ^30^ |
| PD utility (2L+ mapped) | 0.69 | 0.67–0.71 | Beta | ^30^ |
| **Utilities (Xu 2022, Chinese EQ-5D)** |  |  |  |  |
| PFS utility (1L) | 0.617 | 0.494–0.740 | Beta | ^20^ |
| PD utility | 0.37 | 0.296–0.444 | Beta | ^20^ |
| **Adverse-event disutilities** |  |  |  |  |
| Anaemia disutility | −0.119 | 20% SE | Normal | ^20^ |
| Nausea disutility | −0.21 | 20% SE | Normal | ^20^ |
| **Adverse-event rates (PROpel safety population, n=398 int / n=396 ctrl)** |  |  |  |  |
| Grade ≥3 anaemia (intervention) | 16.1% (64/398) | Beta(65,335) | Beta | ^7^ |
| Grade ≥3 anaemia (control) | 3.3% (13/396) | Beta(14,384) | Beta | ^7^ |
| Any-grade nausea (intervention) | 30.7% (122/398) | Beta(123,277) | Beta | ^7^ |
| Any-grade nausea (control) | 14.4% (57/396) | Beta(58,340) | Beta | ^7^ |
| **Costs (2025 CNY, per 28-day cycle)** |  |  |  |  |
| Olaparib (300 mg BID) | 188.7 | 20% SE | Gamma | ^22^ |
| Abiraterone (1000 mg QD) | 717.6 | 20% SE | Gamma | ^23^ |
| Prednisone (5 mg BID) | 3.78 | 20% SE | Gamma | ^20^ |
| PFS management (non-drug) | 2,595.75 | 20% SE | Gamma | ^20^ |
| Post-progression management | 5,831.53 | 20% SE | Gamma | ^20^ |
| Anaemia treatment (one-time) | 3,893.05 | 20% SE | Gamma | ^20^ |
| Nausea treatment (one-time) | 467.49 | 20% SE | Gamma | ^20^ |
| **Analysis parameters** |  |  |  |  |
| Time horizon | 15 years | — | Fixed | Per 99% survival capture |
| Cycle length | 28 days | — | Fixed | PROpel schedule ^6^ |
| Discount rate | 5% | — | Fixed | Chinese PE guidelines ^21,41^ |
| WTP threshold | CNY 287,247/QALY | 95,749–383,996 | Fixed | 3× 2024 per-capita GDP ^24^ |

### Table 2. Base-case cost-utility results — population-specific survival inputs (lognormal PSM)

| Scenario | Total cost int (CNY) | Total QALY int | Total cost ctrl (CNY) | Total QALY ctrl | Incremental cost (CNY) | Incremental QALY | ICER (CNY/QALY) | Cost-effective at CNY 287,247? |
| --- | --- | --- | --- | --- | --- | --- | --- | --- |
| BRCAm × Castro | 348,585 | 5.907 | 154,372 | 2.357 | 194,214 | 3.550 | **54,708** | Yes |
| BRCAm × Xu | 348,585 | 4.586 | 154,372 | 1.730 | 194,214 | 2.856 | 68,000 | Yes |
| ITT × Castro | 228,703 | 3.351 | 207,029 | 2.812 | 21,674 | 0.539 | 40,198 | Yes |
| ITT × Xu | 228,703 | 2.459 | 207,029 | 1.963 | 21,674 | 0.496 | 43,721 | Yes |

Notes: The BRCAm and ITT populations use different control-arm OS construction methods reflecting their difference in data maturity (§2.5). For the BRCAm subgroup (BRCA1/2-confirmed, n=85) the OS control curve is a median-anchored, maturity-constrained lognormal fixed to the published BRCAm control-arm median OS of 23.0 months (meanlog μ = ln 23.0 = 3.1355, sdlog σ = 1.1422, reproducing the observed 25/38 = 65.8% control mortality at the 36.6-month cut-off); the three Guyot-fitted parametric alternatives are not applicable to BRCAm because the BRCAm placebo arm (n=38, 25 OS events) is too sparse for Guyot digitisation (§2.5), and BRCAm parametric uncertainty is quantified probabilistically via the PSA (Table 4). For the ITT population the OS control curve was re-fit in flexsurv v2.3 to Guyot-reconstructed pseudo-IPD from the PROpel Number-at-Risk table (Saad 2023, Lancet Oncol), median 35.0 months (meanlog 3.556, sdlog 1.118). Intervention-arm curves are derived by proportional-hazards scaling with the true BRCAm hazard ratios (rPFS 0.23; OS 0.29). See Supplementary Table S2 for the ITT distributional matrix (Panel B, valid Guyot IPD) and the BRCAm PH-versus-AFT robustness PSA (Panel A).

### Table 3. Partition survival vs Markov structural robustness (base case ICERs)

| Scenario | PSM ICER (CNY/QALY) | Markov ICER (CNY/QALY) | Ratio (Markov / PSM) | Both CE at WTP CNY 287,247? |
| --- | --- | --- | --- | --- |
| BRCAm × Castro | 54,708 | 81,878 | 1.50 | Yes |
| BRCAm × Xu | 68,000 | 120,558 | 1.77 | Yes |
| ITT × Castro | 40,198 | 80,479 | 2.00 | Yes |
| ITT × Xu | 43,721 | 104,927 | 2.40 | Yes |

Note: The co-primary two-transition Markov is reported here. PSM and Markov use the same control-arm parameterisation (BRCAm median-anchored lognormal OS control median 23.0 months, meanlog 3.1355, sdlog 1.1422; ITT Guyot lognormal meanlog 3.5555, sdlog 1.118, median 35.01 months) so that the ratio isolates PSM cycle-loop versus Markov transition-matrix structure from parametric shape change. The Markov PFS→PD and PD→Death hazards are calibrated arm-by-arm so each arm reproduces BOTH its median rPFS and its median OS (BRCAm control 8.0/23.0 mo and intervention 34.8/79.3 mo under a prespecified naive median-ratio calibration using HR_PFS 0.23 and HR_OS 0.29 (these are Markov targets, not observed or PSM PH-scaled medians); ITT control 19.8/35.0 mo, intervention 30.0/43.2 mo). Under these BRCAm calibration targets the intervention OS target (79.3 months = 23.0/0.29) far exceeds the rPFS target (34.8 months), so the intervention arm accrues a long, costly post-progression occupancy; this makes the two-transition Markov the HIGHEST of the three structures examined for BRCAm (Markov/PSM ratio 1.50–1.77) — the reverse of the HRRm-era ordering. A single-transition Markov reproduces the ITT results but is undefined for the BRCAm intervention arm, because no non-negative single PD→Death hazard can reproduce an OS median of 79.3 months from the PH-scaled rPFS curve (zero-hazard boundary); the two-transition calibration is therefore used as the co-primary Markov specification (§3.2). Adverse-event cost and disutility are applied one-off at cycle 0, matching the PSM convention. The maximum ICER across all structures and scenarios is CNY 120,558/QALY (BRCAm × Xu, two-transition Markov), 42.0% of the WTP threshold, so the cost-effectiveness conclusion is invariant to structural specification.

### Table 4. Probabilistic sensitivity analysis (5,000 iterations, seed 20260707)

| Scenario | Mean ICER | Median ICER | 95% UI (lower) | 95% UI (upper) | P(CE @ 95,749) | P(CE @ 191,498) | P(CE @ 287,247) | P(Dominance) |
| --- | --- | --- | --- | --- | --- | --- | --- | --- |
| BRCAm × Castro | 57,156 | 56,408 | 37,420 | 81,808 | 0.997 | 1.000 | 1.000 | 0.000 |
| BRCAm × Xu | 72,756 | 70,887 | 44,747 | 111,424 | 0.904 | 1.000 | 1.000 | 0.000 |
| ITT × Castro | 15,891 | 44,993 | -162,188 | 93,315 | 0.984 | 0.999 | 0.999 | 0.194 |
| ITT × Xu | 37,574 | 47,626 | -106,526 | 130,182 | 0.870 | 0.999 | 1.000 | 0.199 |

Notes: ICER units CNY/QALY. WTP thresholds: 1× GDP = CNY 95,749; 2× GDP = CNY 191,498; 3× GDP = CNY 287,247. P(CE) is net-monetary-benefit-based (proportion with NMB > 0). P(Dominance) is the proportion of PSA iterations with positive incremental QALYs and negative incremental cost (favourable outcome).

### Table 5. EVPI and EVPPI at peak-EVPI WTP by scenario

| Scenario | Peak-EVPI WTP (CNY/QALY) | Per-patient EVPI (CNY) | EVPPI: HRs (%) | EVPPI: Utilities (%) | EVPPI: PFS mgmt cost (%) | EVPPI: PD cost (%) | EVPPI: Drug costs (%) | EVPPI: AE parameters (%) |
| --- | --- | --- | --- | --- | --- | --- | --- | --- |
| BRCAm × Castro | 57,000 | 16,002 | 5,477 (34%) | 4,741 (30%) | 13,330 (83%) | 3,632 (23%) | 4,341 (27%) | 2,439 (15%) |
| BRCAm × Xu | 72,000 | 18,139 | 6,877 (38%) | 8,282 (46%) | 13,012 (72%) | 3,622 (20%) | 4,121 (23%) | 2,196 (12%) |
| ITT × Castro | 41,000 | 8,351 | 6,922 (83%) | 1,467 (18%) | 3,367 (40%) | 3,021 (36%) | 1,660 (20%) | 1,059 (13%) |
| ITT × Xu | 44,000 | 9,748 | 8,419 (86%) | 1,841 (19%) | 3,414 (35%) | 3,110 (32%) | 1,770 (18%) | 1,190 (12%) |

Notes: Per-patient EVPPI values (CNY) estimated by nonparametric random-forest regression per Strong et al. 2014 ^46^ using min_samples_leaf = 20 (Strong 2014 default, matching the specification in Methods §2.6). Percentages express EVPPI as a fraction of the scenario EVPI at peak-EVPI WTP; parameter subsets are non-orthogonal (e.g., cost and survival covariates jointly determine incremental cost), so EVPPI subset values do not sum to 100%. At the CNY 287,247/QALY WTP threshold, EVPI is zero for all scenarios and EVPPI is not informative. Table 5 subset shares are drawn from the primary EVPPI run (RF min_samples_leaf = 20, subset-share sums 203% to 212%); the independent OOB / GAM metamodel comparison on the same n=5000 seed-20260707 PSA draws is reported in Supplementary Table S1 (GAM subset sums 172% to 193%, RF-OOB subset sums 199% to 215%). Rank preservation of dominant subsets across RF, OOB, and GAM is confirmed for all four scenarios (PFS management cost dominates both BRCAm scenarios; hazard ratios dominate both ITT scenarios). Peak-EVPI WTP shifts upward for the BRCAm scenarios relative to the previous parameterisation (49,000→57,000 Castro; 59,000→72,000 Xu) because the corrected true-BRCAm base ICER is higher; ITT peak-EVPI WTP is unchanged.

### Table 6. Five-year budget impact (CNY, Chinese healthcare payer perspective)

| Uptake scenario | Year 2025 | Year 2026 | Year 2027 | Year 2028 | Year 2029 | 5-year cumulative |
| --- | --- | --- | --- | --- | --- | --- |
| Low (10% to 30%) | 193,398 | 1,174,667 | 3,297,595 | 6,924,930 | 12,434,954 | **24,025,544** |
| Base (20% to 50%) | 386,796 | 2,349,335 | 6,595,190 | 13,650,602 | 23,592,563 | **46,574,486** |
| High (40% to 70%) | 773,592 | 4,438,227 | 11,476,971 | 22,216,569 | 36,625,367 | **75,530,727** |

Probabilistic (10,000 Monte Carlo iterations, seed 20260717; all six BIA cascade parameters propagated jointly): mean 5-year cumulative net budget impact CNY 21.88 million (95% CI 13.16–33.36) for low uptake, CNY 42.42 million (25.51–64.67) for base uptake, and CNY 68.79 million (41.37–104.88) for high uptake; the pooled 95% interval across the combined uptake-scenario draws is CNY 14.9–94.0 million (Figure 8), distinct from the union of scenario-specific intervals.

Notes: Budget impact = intervention annual cost − control-arm annual cost, computed on the true-BRCAm OS HR of 0.29 (§3.6). The deterministic 5-year net budget impact rises about five-fold relative to the v7.7.x HRRm-mislabeled estimate (base CNY 9.41 million at the incorrect HR 0.66) because the corrected strong OS benefit keeps a substantial fraction of intervention patients alive and on therapy within the undiscounted 5-year window; the corrected magnitude returns to the level implied by the interim BRCAm HR (about 0.30). Cumulative counterfactual all-comer cost over 5 years (all BRCAm-eligible patients on control) is CNY 330,096,174 and is unchanged by the correction because the control arm is invariant. Tentative operational assumptions: BRCA testing rate ramping 30% to 70% over 2025 to 2029; 60% of Chinese mCRPC patients eligible for first-line combination therapy. The BIA cascade applies a 10% BRCA1/2 prevalence based on Western trial-population estimates ^7,16^; under a lower Chinese-specific germline prevalence of 5.6% ^5^, the 5-year cumulative BIA scales proportionally to CNY 13.46 million (low), CNY 26.08 million (base), and CNY 42.30 million (high uptake). This proportional scaling is an approximation because the 10% Western figure derives from tumour-based BRCA1/2 testing (germline plus somatic) as used in PROpel eligibility, whereas the 5.6% Chinese figure derives from germline-only testing in an unselected prostate cancer cohort ^5^. Chinese tumour-based testing would additionally identify somatic BRCA1/2 alterations not captured by germline-only assays, so the 5.6% figure is a lower bound on the Chinese tumour-testing-positive rate rather than a direct equivalent of the 10% Western tumour-testing figure. Because the qualitative reimbursement conclusion depends on affordability at the payer level rather than absolute magnitude, the BIA range under either prevalence assumption remains within the range of routine national reimbursement decisions for oncology combination therapies in China.
